# Fitness and Exercise Effects on Brain Age: A Randomized Clinical Trial

**DOI:** 10.1101/2025.02.25.25322645

**Authors:** Lu Wan, Cristina Molina-Hidalgo, Mary E. Crisafio, George Grove, Regina L. Leckie, Thomas W. Kamarck, Chaeryon Kang, Mia DeCataldo, Anna L. Marsland, Matthew F. Muldoon, Mark R. Scudder, Javier Rasero, Peter J. Gianaros, Kirk I. Erickson

## Abstract

**Objectives:** Examine the effect of aerobic exercise on structural brain age and explore potential mediators.

**Methods:** In a single-blind, 12-month randomized clinical trial, 130 healthy participants aged 26-58 years were randomized into a moderator-to-vigorous intensity aerobic exercise group or a usual-care control group. The exercise group attended 2 supervised 60-minute sessions per week in a laboratory setting plus home-based exercise to achieve 150 minutes of exercise per week. Brain-predicted age difference (brain-PAD) and cardiorespiratory fitness (CRF) were assessed at baseline and 12 months. Intention-to-treat (ITT) and completers analyses were performed.

**Results:** The 130 participants (67.7% female) had a mean (SD) age of 41.28 (9.93) years. At baseline, higher CRF (VO_2peak_) was associated with smaller brain-PAD (β=-0.309, p=0.012). After the intervention, the exercise group showed a decrease in brain-PAD (estimated mean difference (EMD) =-0.60; 95% CI: -1.15 to -0.04; p=0.034) compared to the control group (EMD=0.35; 95% CI: -0.21 to 0.92; p=0.22); time×group interaction (between-group difference (BGD)= -0.95; 95% CI: -1.72 to -0.17; p=0.019). VO_2peak_ improved in the exercise group (EMD=1.60; 95% CI: 0.29 to 2.90; p=0.017) compared to the control group (EMD=-0.78; 95% CI: -2.17 to 0.60; p=0.26); time×group interaction (BGD=2.38; 95% CI: 0.52 to 4.25; p=0.015). Body composition, blood pressure, and brain-derived neurotrophic factor levels were unaffected. None of the proposed pathways statistically mediated the effect of exercise on brain-PAD. The results from completers were similar.

**Conclusion:** Engaging in 12 months of moderate-to-vigorous exercise reduced brain-PAD in early-to-midlife adults. The pathways by which these effects occur remain unknown.

**Summary Box:** *What is already known on this topic:* Midlife risk factors influence brain aging, with physical activity conferring protective benefits, yet evidence for the effect of exercise on midlife brain age and underlying mechanisms remains limited.

*What this study adds:* Participation in a 12-month aerobic exercise intervention significantly reduced a neuroimaging marker of brain age. Higher cardiorespiratory fitness was also associated with younger brain age.

*How this study might affect research, practice or policy:* Findings of this study complement the scarce literature examining the effects of exercise on early-to-midlife brain health and confirm the neuroprotective effects of aerobic exercise against accelerated brain aging in early-to-midlife adults.

## INTRODUCTION

Early adulthood through midlife is a dynamic period in which risk factors for age-related brain atrophy, deterioration, and dementia can be modified by lifestyle behaviors.^1–4^ For example, midlife hypertension and obesity confer risks for late-life dementia, while greater physical activity (PA) protects against cognitive decline and Alzheimer’s disease (AD) pathology in late adulthood.^5,6^ Observational studies suggest a cumulative and protracted neural impact of cardiometabolic health and lifestyle that begins decades before clinical manifestation of age-related cognitive impairment.^3^ Despite the importance of midlife risk factors, most PA interventions designed to improve cardiorespiratory fitness (CRF) and examine emergent brain health outcomes have been limited to late adulthood.^7–10^ Moreover, such studies have focused on the morphology and function of specific rather than global brain health or brain aging indicators, such as the volume of the hippocampus and prefrontal cortex. In these regards, there remains a need to clarify whether exercise in early and mid-adulthood could modify the trajectory of biomarkers that reliably reflect aging-related brain health.

Brain-predicted age difference (*brain-PAD*), derived from structural magnetic resonance imaging (MRI) using machine learning algorithms, has emerged as a sensitive biomarker of brain health. Brain-PAD quantifies the gap between chronological age and predicted *brain age*, interpreted as a surrogate biomarker indicating how much “older” or “younger” a given brain appears relative to the person’s chronological age. The machine learning approaches used to estimate *brain age* are more sensitive than traditional techniques for detecting small differences in brain morphology, which is particularly useful when studying an age range in which individual variation in brain structure is more subtle than in later life.^11^ While cross-sectional evaluations of brain age cannot be used to infer rates of aging *per se*,^12^ greater brain-PAD values predict future poorer cognitive performance,^13,14^ earlier mortality,^15^ and accelerated neurocognitive decline and dementia, including progression from mild cognitive impairment to AD ^16,17^ and other dementias.^12,18^ Therefore, *brain-PAD* may be capturing several important attributes of brain health that are predictive of long-term clinical endpoints.

In addition, the biological and physiological factors that mediate the effects of exercise on brain health remain poorly understood.^19,20^ Several studies suggest that increases in CRF might mediate improvements in brain health. In fact, higher CRF is associated with elevated cognitive performance and reduced risk for dementia,^21,22^ and interventions that modify CRF are effective at offsetting age-related declines in brain morphology.^23,24^ In addition, exercise-induced decreases in cardiometabolic risk factors, such as blood pressure, have also been identified as candidate pathways because sustained elevations in blood pressure have negative consequences for brain health.^25^ Other studies suggest that exercise-induced changes in body composition could alter various signaling and inflammatory pathways, thereby leading to improved brain health.^26,27^ Finally, exercise-induced changes in brain-derived neurotrophic factor (BDNF) have been speculated to drive changes in brain morphology.^23^ Yet, despite this speculation, there is limited evidence in support of these pathways in humans.

In this study, our objectives were to evaluate (1) the cross-sectional association between CRF and brain age and (2) the effects of a 12-month aerobic exercise intervention on brain age. We hypothesized that higher CRF would be associated with a ‘younger’ brain age (reduced *brain-PAD*) and that a 12-month aerobic exercise intervention would reduce brain-PAD in those randomly assigned to engage in moderate-to-vigorous aerobic exercise. We also explored several possible mediators of the exercise intervention on brain age, including intervention-induced changes in CRF, body mass, blood pressure, and BDNF.

## METHODS

The trial protocol and statistical analysis plan are provided in Supplement 1. Methodological details are in the Supplemental Methods in Supplement 2.

### Study Design and Participants

Adults aged 26-58 years were recruited for participation in this single-center, parallel-arm RCT (Exercise, Brain, and Cardiovascular Health (eBACH)) (ClinicalTrials.gov: NCT03841669). The study protocol and informed consent were approved by the University of Pittsburgh Institutional Review Board (IRB ID: 19020218). Participants provided written informed consent prior to data collection. The Consolidated Standards of Reporting Trials (CONSORT) reporting guideline was followed.^28^ Participants were eligible if they reported no history or presence of neurological disorders or current use of prescribed blood pressure medication, and exercised for fewer than 100 minutes per week. More details are available regarding participant eligibility and study design in the published protocol.^29^

### Patient and Public Involvement

This was a community-based sample without patient involvement.

### Equity, Diversity, and Inclusion Statement

Recruitment was based on the racial and ethnic diversity in the community. Participants self-identified their race/ethnicity consistent with NIH guidelines.

### Randomization and Blinding

Participants were randomly assigned to either a moderate-to-vigorous intensity aerobic exercise condition or to a health information control condition for 12 months. Randomization was conducted using stratified block randomization on a web-based system through the Research Electronic Data Capture (REDCap) randomization module by the study biostatistician (CK) to ensure unbiased allocation of participants to the two intervention groups by age and sex. All investigators and staff involved in data collection were blinded to group assignment. Only staff involved in the implementation of the exercise intervention, scheduling of sessions, and study coordination were unblinded to group assignment.

### Exercise Intervention and Control

The exercise group was instructed to attend two supervised 60-minute exercise sessions per week in a laboratory at the University of Pittsburgh, along with home-based exercise, to achieve the prescribed 150 minutes of exercise per week. Participants were encouraged to walk, jog, or run on a treadmill, as well as to record the use of other types of aerobic exercise equipment such as bikes, elliptical machines, stair climbers, and rowers. Participants assigned to the health information control group were asked not to change their exercise patterns. They were provided information about the health benefits of engaging in PA and were contacted approximately every 6 weeks for continued contact with the investigative team.

### Outcome Measurements

#### Brain-PAD

The MRI acquisition and preprocessing steps are detailed in Supplement 2. Brain age estimation was performed on T1-weighted images using brainageR (v2.1), an open-access software for generating brain-predicted age (github.com/james-cole/brainageR).^30^ BrainageR was pre-trained to predict brain age from normalised brain volumetric maps of 3377 healthy adults (aged 18-92 years).^31^ The brainageR model was applied to the preprocessed T1-weighted images in the current study to estimate a brain-predicted age score for each participant at each time point. Brain-PAD was calculated as the deviations of predicted brain age from chronological age.

#### Cardiorespiratory Fitness

CRF was measured with a graded exercise test (GXT) using a modified Balke Protocol ^32^ at both baseline and 12-month assessments. Peak oxygen uptake (VO_2peak_) was the highest VO_2_ value obtained during the maximal test and represents the measure of CRF used here.

#### Biological Measures

Body mass index (BMI, kg/m^2^) was calculated by dividing weight (kg) and height (m^2^) recorded before the GXT. Body composition, including waist circumference (WC), height, weight, and percentage of body fat, were measured at both baseline and follow-up. Resting systolic blood pressure (SBP) and diastolic blood pressure (DBP) were computed from seated resting blood pressures obtained with an Omron IntelliSense© BP Monitor. Mean arterial pressure (MAP) was further calculated from SBP and DBP. Plasma levels of BDNF were measured from fasting blood samples. More information about the blood assays is provided in Supplement 2.

### Power and Sample Size

We were powered to detect effect sizes between 0.5 and 0.6 (Cohen’s d) for changes in brain volume and cardiovascular disease biomarkers resulting from the exercise intervention. 60 participants per group were deemed sufficient to test our hypotheses with an α error of 5% at 80% power, after accounting for an estimated 20% attrition rate.

### Statistical Analysis

Multiple linear regression was used to investigate the association between CRF and brain-PAD at baseline, adjusting for chronological age, sex, years of education, and BMI. Image quality rating (IQR) generated from CAT12 was also included as a covariate ^33^. Statistical inferences were conducted at the significance level of 0.05 (p<0.05).

We assessed the effect of the exercise intervention on brain-PAD, CRF, and other biological measures using linear mixed modeling (LMM). The LMM models included the fixed effect for treatment (12-month aerobic exercise, control), time (baseline, month 12), and their interaction. We also included a random intercept for individual participants to account for the within-individual correlation among repeated measures at baseline and 12 months. For brain-PAD, we adjusted for baseline chronological age, sex, years of education, BMI, and IQR. For CRF, we controlled for age, sex, years of education, and BMI. The LMM considers all available data, which conforms to the intention-to-treat (ITT) analytical framework. A secondary analysis of intervention completers was conducted.

A prerequisite for testing mediation is that the changes in outcome must be correlated with the changes in the mediator. As such, we tested the relationship between changes in brain-PAD and changes in CRF, body mass, blood pressure, and BDNF with multiple linear regression models, controlling for chronological age, sex, and years of education. We also assessed associations by collapsing across both groups and solely in the aerobic exercise group. Unstandardized B-values, standardized β-values, and p-values were reported.

All analyses were performed using R software (Version 4.2.1). Data were analyzed from November 2023 to June 2024.

## RESULTS

The participant flow diagram is presented in Figure 1. Participants were recruited from May 2019 to October 2022 and followed up through February 2024. In total, 811 participants were assessed for eligibility, 130 were randomized equally into the aerobic PA and control conditions, and 81 (62.3%) completed post-intervention assessments. Additional details about missing data are in Supplement 2.

**Figure 1.**
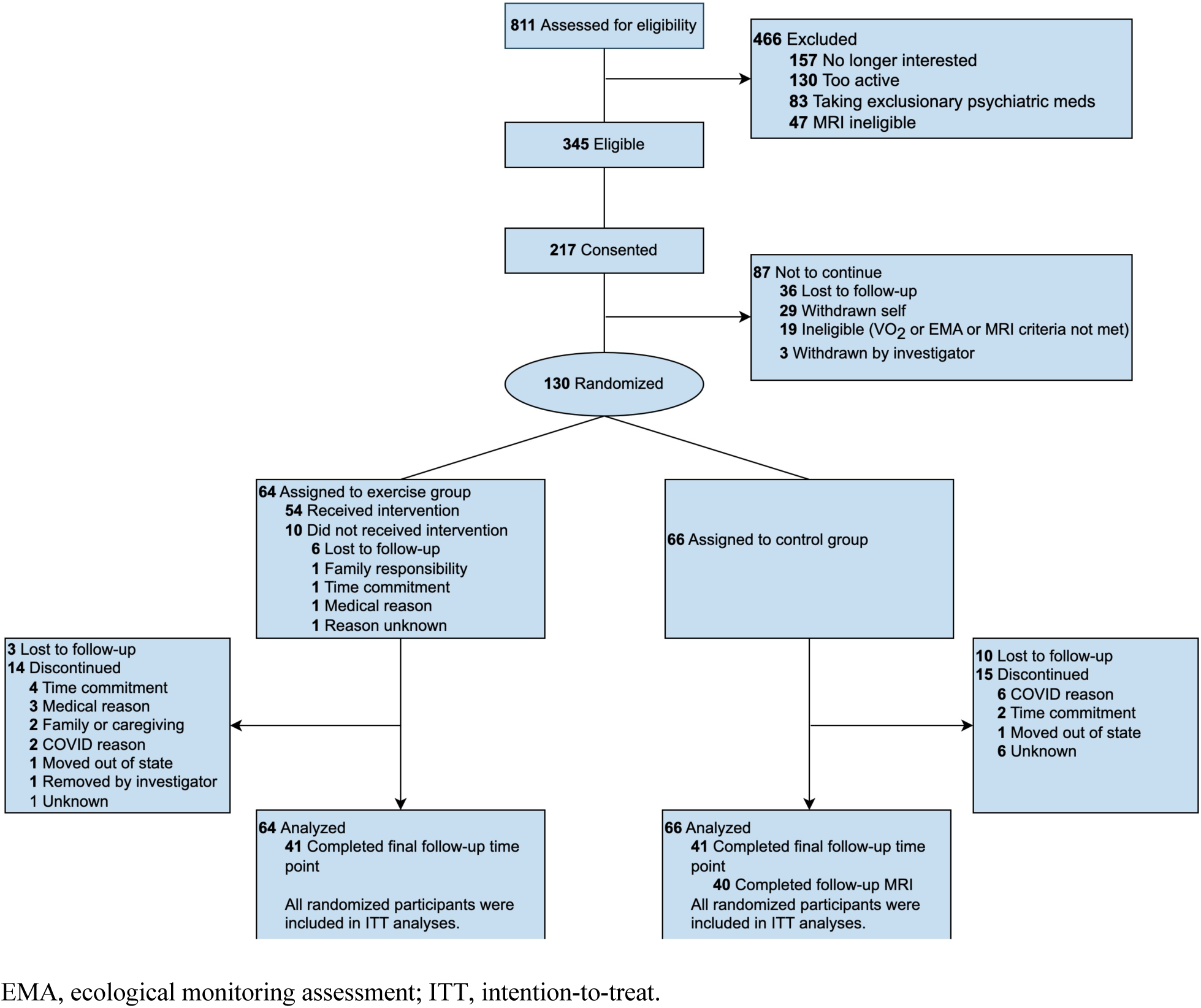
CONSORT Flowchart of Sample Inclusion.

### Participants

Demographic characteristics are described in Table 1. Baseline demographic characteristics were balanced between groups; however, there was a significant difference in baseline VO_2peak_ between control and exercise groups (B=2.435, p=0.005), after adjusting for sex, age, years of education, and BMI.

**Table 1.** Baseline Characteristics by Study Arm.

| Variable | 150min AEx | Control | Total Group | p-value<sup>a</sup> |
| --- | --- | --- | --- | --- |
| Number, n | 64 | 66 | 130 |  |
| Age, mean (SD), y | 41.55 (10.48) | 41.03 (9.43) | 41.28 (9.93) | 0.856 |
| Sex |  |  |  | 0.904 |
| Female, No. (%) | 43 (67.2) | 45 (68.2) | 88 (67.7) |  |
| Male, No. (%) | 21 (32.8) | 21 (31.8) | 42 (32.3) |  |
| Race |  |  |  | 0.350 |
| African American / Black, No. (%) | 11 (17.2) | 5 (7.6) | 16 (12.3) |  |
| Caucasian / White, No. (%) | 42 (65.6) | 49 (74.2) | 91 (70.0) |  |
| Asian, No. (%) | 5 (7.8) | 9 (13.6) | 14 (10.8) |  |
| All Others, No. (%) | 6 (9.4) | 3 (4.6) | 9 (6.9) |  |
| Ethnicity |  |  |  | 0.270 |
| Hispanic or Latinx, No. (%) | 5 (7.8) | 2 (3.0) | 7 (5.4) |  |
| Not Hispanic or Latinx, No. (%) | 59 (92.2) | 64 (97.0) | 123 (94.6) |  |
| Education, mean (SD), y | 17.16 (3.11) | 17.30 (2.82) | 17.23 (2.95) | 0.341 |
| BMI, mean (SD), kg/m<sup>2</sup> | 30.24 (6.66) | 28.66 (6.44) | 29.44 (6.57) | 0.143 |
| VO<sub>2peak</sub>, mean (SD), mL/kg/min | 26.85 (6.60) | 30.23 (6.93) | 28.57 (6.95) | **0.011** |
| Brain-predicted age, y | 42.22 (9.46) | 42.22 (10.54) | 42.22 (9.99) | 0.912 |
| Brain-PAD, y | 0.73 (5.28) | 1.19 (6.53) | 0.97 (5.93) | 0.577 |

<sup>a</sup> Baseline differences between groups were determined using the two-samples Wilcoxon rank sum test for continuous variables and Chi-square test for categorical variables. Statistically significant values at p<0.05 are shown in bold.

Adherence for the exercise intervention group (n=40), evaluated by the ratio of total prescribed minutes exercised, was 93%, with an average of 138.9 minutes of exercise per week accounting for supervised and home-based sessions. Attendance to the supervised exercise sessions was 73%. The average exercise intensity was 124% of the 60% heart rate reserve, with an average RPE of 12.91.

### Cross-sectional Relationship between CRF and Brain-PAD

A graphical illustration of brain-PAD and the age bias ^34,35^ is presented in Figure S1 and Supplement 2. At baseline, higher CRF was associated with lower brain-PAD (β=-0.309, p=0.012; Figure 2), controlling for covariates. For every 1SD increase in VO_2peak_ (about 7 mL/kg/min), brain age decreased by about 1.83 years. There was no significant main effect of sex, years of education, or BMI on brain-PAD (all p>0.05; Table S1).

**Figure 2.**
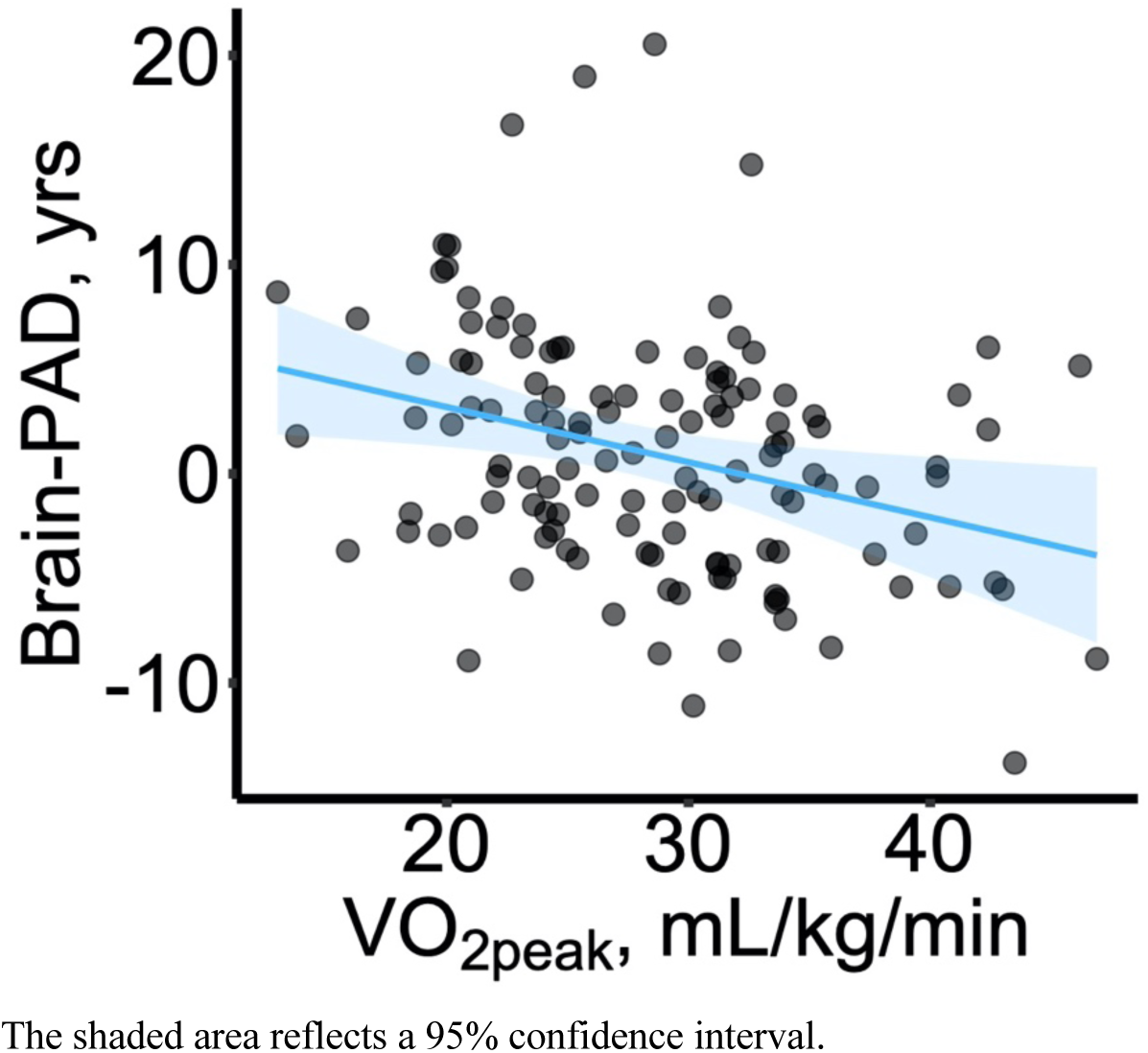
The Cross-sectional Relationship Between CRF and Brain-PAD in the Entire Sample at Baseline.

### Effect of the Exercise Intervention on Brain-PAD

Consistent with our predictions, the exercise intervention group showed decreased brain-PAD at the 12-month follow-up compared to baseline (-0.60 years, 95% CI: -1.15 to -0.04, p=0.034). The control group did not show a significant change in brain-PAD from baseline to 12-months (0.35 years, 95% CI: - 0.21 to 0.92, p=0.217) (see Table 2).

**Table 2.** Estimated Marginal Means for Brain-PAD, CRF, and Biological Measurements Comparing the Intervention and Control Arms.

| Variable | 150min AEx | Control | Difference between arms <sup>b</sup> |  |
| --- | --- | --- | --- | --- |
|  | mean (SE) [95% CI]<sup>a</sup> | mean (SE) [95% CI]<sup>a</sup> | mean (SE) [95% CI] | p |
| Brain-PAD, y | -0.60 (0.28) [-1.15, -0.04] | 0.35 (0.29) [-0.21, 0.92] | -0.95 (0.40) [-1.72, -0.17] | **0.019** |
| *CRF* |  |  |  |  |
| $VO_{2peak}$, mL/kg/min | 1.60 (0.66) [0.29, 2.90] | -0.78 (0.70) [-2.17, 0.60] | 2.38 (0.96) [0.52, 4.25] | **0.015** |
| *Body composition* |  |  |  |  |
| BMI, kg/m<sup>2</sup> | 0.30 (0.20) [-0.09, 0.69] | 0.48 (0.20) [0.08, 0.89] | -0.19 (0.28) [-0.74, 0.37] | 0.515 |
| Body fat, % | 0.14 (0.41) [-0.68, 0.96] | 0.54 (0.43) [-0.31, 1.39] | -0.40 (0.59) [-1.56, 0.76] | 0.497 |
| WC, cm | 0.47 (0.92) [-1.37, 2.31] | 2.37 (0.96) [0.46, 4.29] | -1.91 (1.33) [-4.51, 0.70] | 0.156 |
| *Blood pressure* |  |  |  |  |
| SBP, mm Hg | -2.02 (1.71) [-5.43, 1.38] | 1.24 (1.76) [-2.26, 4.75] | -3.27 (2.45) [-8.01, 1.58] | 0.186 |
| DBP, mm Hg | -0.82 (1.40) [-3.61, 1.96] | -0.70 (1.44) [-3.57, 2.16] | -0.12 (2.00) [-3.99, 3.87] | 0.952 |
| MAP, mm Hg | -1.44 (1.43) [-4.29, 1.41] | -0.01 (1.47) [-2.94, 2.92] | -1.43 (2.05) [-5.39, 2.64] | 0.487 |
| *Blood biomarker* |  |  |  |  |
| log BDNF, pg/ml | 0.35 (0.14) [0.07, 0.62] | -0.04 (0.14) [-0.32, 0.24] | 0.39 (0.20) [0.01, 0.78] | 0.053 |

<sup>a</sup> Differences were calculated as values at 12 months minus baseline.
<sup>b</sup> Differences between arms were calculated as 150min AEx group minus control group.
Abbreviations: Brain-PAD, brain-predicted age difference; CRF, cardiorespiratory fitness; BMI, body mass index; WC, waist circumference; SBP, systolic blood pressure; DBP, diastolic blood pressure; MAP, mean arterial pressure; BDNF, brain-derived neurotrophic factor.

We found a significant Time × Group interaction such that the exercise intervention group showed a significant decrease in brain-PAD from baseline to 12 months compared to the control group with a marginal mean difference of -0.95 (95% CI: -1.72 to -0.17, p=0.019), adjusting for covariates (Table 2; Figure 3A). The intervention effect on brain-PAD remained after further controlling for baseline differences in VO_2peak_. Analyses on completers yielded similar results (Figure S2A, Table S3).

**Figure 3.**
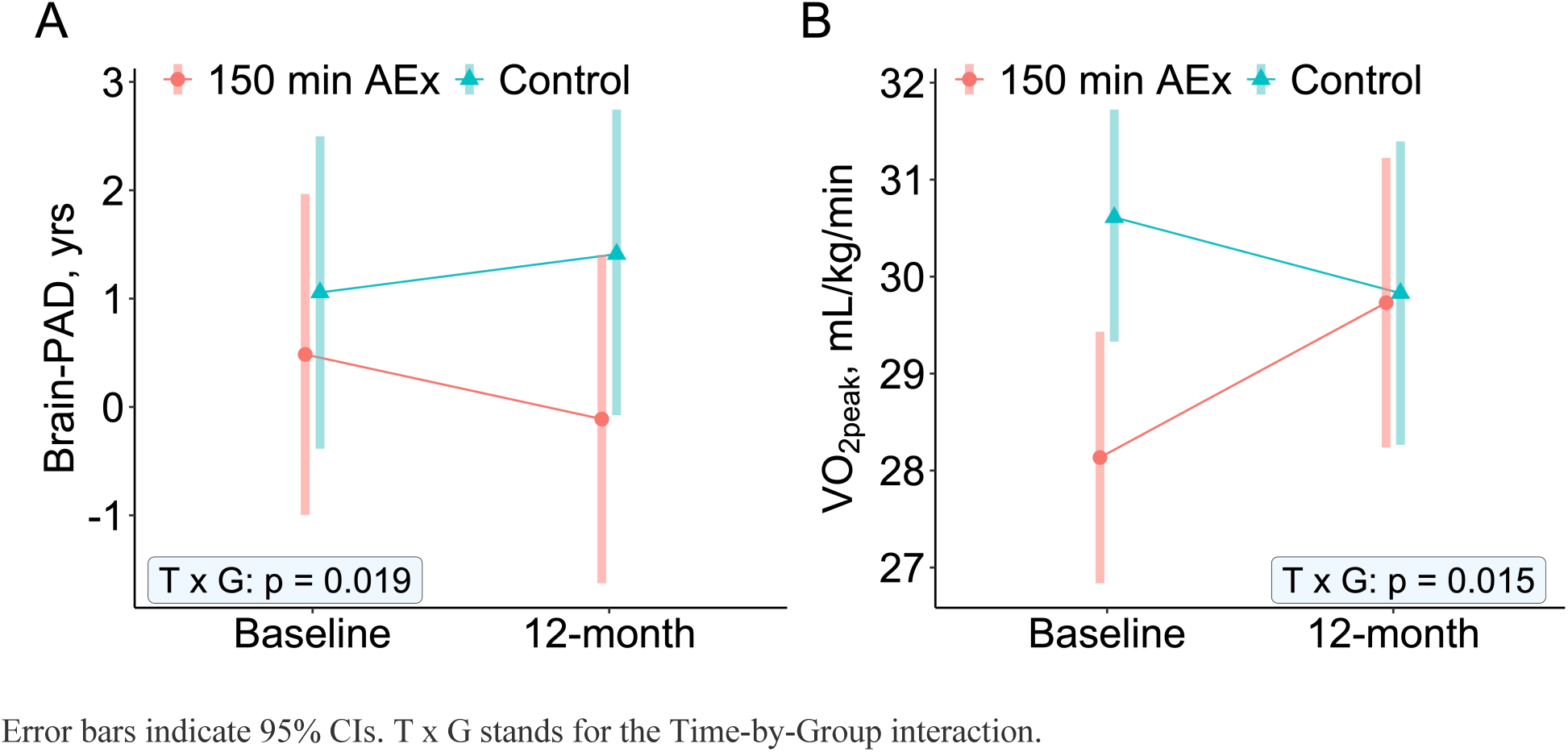
Intervention Effect with Changes in Marginal Mean Values of (A) Brain-PAD and (B) VO_2peak_ from Baseline to 12 Months by ITT Allocation.

### Effect of the Exercise Intervention on CRF

The aerobic exercise group showed significant improvement in VO_2peak_ at 12 months compared to baseline (1.60 mL/kg/min, 95% CI: 0.29 to 2.90, p=0.017), whereas the control group had a slightly decreased VO_2peak_ from baseline to 12-months (-0.78 mL/kg/min, 95% CI: -2.17 to 0.60, p=0.265) (see Table 2).

A significant Time × Group interaction was found for VO_2peak_ such that there was an increase from baseline to 12 months in the exercise intervention group compared to controls with a marginal mean difference of 2.38 (95% CI: 0.52 to 4.25, p=0.015), adjusting for covariates (Table 2). Changes in VO_2peak_ by group assignment are shown in Figure 3B. Analyses on completers also yielded similar results (Figure S2B, Table S3).

### Associations between Changes in Brain-PAD, CRF, and Other Biological Measures

We explored possible physiological and biological mediators that may explain how the exercise intervention led to a decrease in brain-PAD. We found no statistical associations between changes in VO_2peak_ and changes in brain-PAD when examining the entire sample, or when stratifying by intervention group (all p>0.05; Figure 4; Figure S3). There was a non-significant trend that greater improvements in CRF were associated with more substantial decreases in brain-PAD in the aerobic exercise group (β=- 0.244, t=-1.473, p=0.15; Figure S3B).

**Figure 4.**
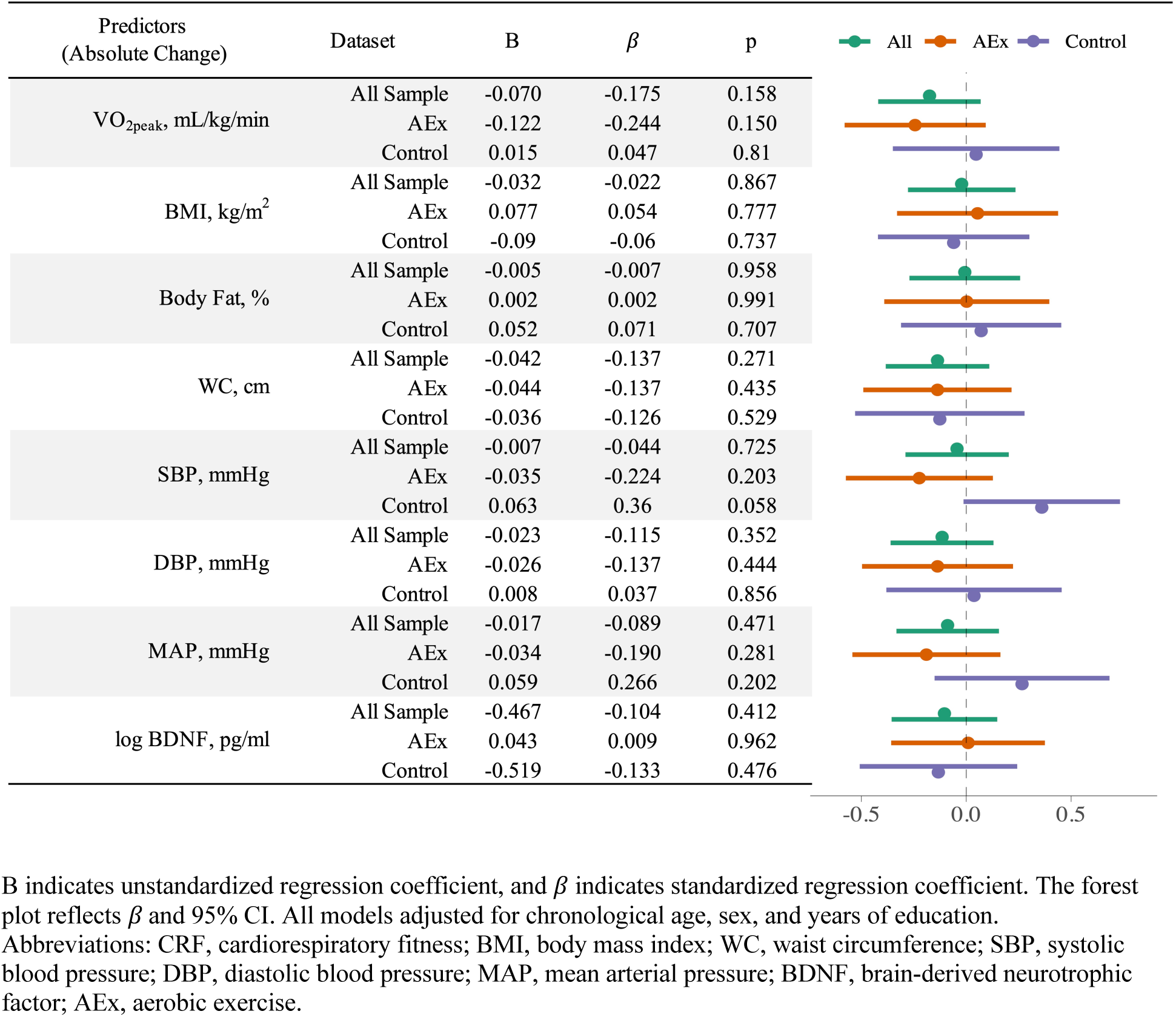
Association between Changes in Brain-PAD and Changes in CRF, Metrics of Body Composition, Blood Pressure, and BDNF.

The exercise intervention did not modify body composition or blood pressure, as demonstrated by non-significant Time × Group interactions (all p>0.05; Table 2). There was a marginally significant Time × Group interaction for BDNF such that the exercise group demonstrated greater increases in plasma BDNF levels compared to the controls (95% CI: 0.005 to 0.776, p=0.053; Table 2). However, there were no significant associations between changes in brain-PAD and changes in body composition, blood pressure, or BDNF in the entire sample or the aerobic exercise group (all p>0.05; Figure 4). This suggests that none of these possible biological pathways mediated the effect of the intervention on brain age.

### Adverse Events

A total of 7 adverse events were reported (Table S4) with no causal relationships to the exercise intervention. In the aerobic exercise group, 1 participant had a serious adverse event related to tumor removal, 5 reported musculoskeletal issues, and 1 had a cardiovascular-related event.

## DISCUSSION

We predicted that higher CRF would be associated with lower brain age, and that participation in a 12-month exercise intervention would decrease brain age. We also explored potential biological mediators of the exercise intervention. Consistent with our predictions, higher CRF was associated with younger brain age, and participation in the 12-month exercise intervention significantly reduced brain age. These findings indicate that a structured 12-month aerobic exercise program could promote a “younger” appearing brain based on MRI features of brain structure.

Every standard deviation increase in VO_2peak_ (about 7 mL/kg/min) was associated with approximately 1.83 years decrease in brain-PAD. These results suggest that elevated fitness levels may make the brain less vulnerable to midlife aging, even after adjusting for other confounders. This is consistent with the literature in which self-reported physical activity was associated with younger brain-predicted age ^36,37^ and evidence linking CRF with gray matter volume and white matter integrity ^38–40^. However, caution is warranted when interpreting cross-sectional data -- as a positive brain-PAD is more strongly associated with early life influences as compared to behaviors during middle and older adulthood. In contrast, results from an intervention eliminate some of these challenges to interpretation.

Our analyses demonstrated that the 12-month exercise intervention decelerated brain aging. After 12 months, the brain-PAD of participants in the exercise group decreased by an average of 0.6 years. Note that in recent studies of brain age, every additional year of brain-PAD incurred a 3% relative increased risk of a future dementia diagnosis.^31^ To our knowledge, the current study provides novel evidence that brain age can be reversed by engaging in moderate-to-vigorous physical activity over a 12-month period. Other studies have focused on the effect of aerobic exercise on regional brain volumes or thickness (e.g., hippocampal volume ^41^, frontal gray matter volume and cortical thickness ^42,43^), but few have examined a brain age variable that compares the volumetric information to that of a normative sample. In contrast to our results, a recent study reported no significant changes in brain age after a 6-month exercise intervention in a cohort of cognitively normal older adults aged 65 to 84 years.^44^ Such inconsistency could be related to exercise intensity, duration, and individual characteristics of participants. There also might be greater benefits of a longer exercise intervention in midlife prior to more accelerated brain aging in later life. However, these explanations remain speculative, as there is no study that investigated dose-response relationships between training duration and changes in brain structure in younger or midlife adults.^19^

To explore possible biological mediators of intervention-induced change in brain age, we considered changes in CRF, body composition, blood pressure, and BDNF. We did not find evidence that any of these proposed pathways mediated the impact of the intervention on brain-PAD. It is possible that other pathways, such as inflammatory, mitochondrial, and metabolic pathways, could explain the reduction in brain age induced by the intervention. Future research is warranted to test alternative pathways.

This study has several strengths. First, we focused on early to midlife, a period that has received relatively little attention in the context of exercise and brain health.^19,45^ and provide novel evidence that aerobic exercise influences structural markers of brain health during a critical period of the lifespan.

Second, the brain age measure allowed an assessment of estimated biological brain age compared to chronological age. Third, VO_2peak_ provided an objective, gold-standard index of CRF. Finally, the randomized controlled trial helps to elucidate the causal relationship between CRF, exercise, and brain health.

### Limitations

Limitations of the study include a relatively small sample size, which reduced statistical power to explore possible moderators. Next, the study was influenced by the COVID-19 pandemic. Many participants were hampered during the pandemic by illness, family and child-rearing limitations, or professional challenges that heightened the attrition rate. However, despite the challenges of conducting an exercise intervention in the context of stay-at-home advisories, our adherence rate remained impressively high. Finally, we did not consider other lifestyle risk factors, including alcohol consumption, smoking, etc., which have been reported to influence brain age.^46,47^

### Clinical Implications

These results highlight the potential of structured aerobic exercise programs as an effective, non-pharmaceutical intervention to slow or even reverse brain aging. Promoting regular moderate-to-vigorous physical activity could be a key strategy in delaying cognitive decline and reducing the risk of neurodegenerative diseases. Notably, the observed decrease in brain-PAD after the exercise intervention suggests that maintaining exercise may provide resilience against age-related brain changes, potentially lowering the long-term risk of dementia. Additional research is necessary to understand the mechanisms of exercise on brain age, which may improve biological targets for altering brain aging.

## Supporting information

Supplement 1_Trial Protocol_Statistical Analysis

## Data Availability

All data produced in the present study are available upon reasonable request to the authors.

## Author Contributions

KIE and LW had full access to all the data in the study and took responsibility for the integrity of the data and the accuracy of the data analysis. KIE, PJG, and LW had a role in the concept and design. MEC, GG, RLL, MRS, LW, CK, and ALM had a role in acquisition, analysis, or interpretation of data. LW, KIE, and PJG drafted the manuscript. LW, CK, and JR did the critical statistical analysis. KIE and PJG obtained funding. CMH, RLL, GG, MEC, ALM, and MRS provided administrative, technical, or material support. All authors critically revised the manuscript for important intellectual content and approved the final version.

## Acknowledgments

We would like to thank all of the participants, staff, faculty, and students who contributed to the eBACH study.

