## Supplement 1_Trial Protocol_Statistical Analysis for "Fitness and Exercise Effects on Brain Age: A Randomized Clinical Trial"

### Exercise, Brain, and Cardiovascular Health (eBACH)

Original Approved Protocol

#### 1.0 **SUMMARY:**

Cumulative epidemiological evidence indicates that physical inactivity confers risk for cardiovascular disease (CVD)[1], and that current physical activity (PA) guidelines (prescribed at >150 minutes per week of moderate to vigorous intensity aerobic PA) can reduce CVD morbidity and mortality[2,3]. The biological and behavioral pathways by which PA may reduce CVD risk is still unclear[4]. In line with the P01 focus on neurobiology of CVD risk, we are conducting a 12-month PA intervention with 150 midlife and inactive adults randomized to: (1) 150 minutes per week of moderate to vigorous intensity exercise (i.e., brisk walking; N=75); or (2) a PA and health information group where we will provide cardiovascular information and active daily monitoring (N=75). A comprehensive battery of cardiorespiratory fitness (CRF), cardiovascular health, physiological biomarkers such as endothelial function, and magnetic resonance imaging (MRI) measures will be assessed at baseline, after 6-months, and again after completion of the intervention.

We have assembled a highly creative, productive, and interdisciplinary team with a long history of collaboration and experience conducting exercise interventions to test the following aims:

**Aim 1**: To determine the *Neurobiology of Exercise and Biological CVD risk factors*:

(1A) *Body-to-Brain hypothesis*: Exercise-induced changes in peripheral markers of CVD risk (i.e., insulin resistance, pulse wave velocity, CRF, oxidative stress, biomarkers of endothelial function) will precede and partly explain (statistically mediate) exercise-induced changes in functional and structural features of some areas within visceral control circuits (prefrontal cortex [PFC], anterior cingulate cortex [ACC], insula, hippocampus, and amygdala).

(1B) *Brain-to-Body hypothesis*: Exercise-induced changes in functional and structural features of some areas within visceral control circuits precede and partly explain (statistically mediate) consequent changes in peripheral autonomic and neuroendocrine mediators of CVD risk that are under neural regulation, including baroreflex sensitivity, heart rate variability, glucocorticoid control.

**Aim 2:** To determine the *Neurobiology of Exercise and Stress- and Affect-related CVD risk factors***:**

(2A) *Stress-related parameters of CVD risk*: Exercise will induce changes in visceral control areas engaged by an fMRI stress battery, and these changes will partly explain exercise-induced reductions in cardiovascular stress reactivity in daily life.

(2B) *Affect-related parameters of CVD risk*: Exercise will induce changes in visceral control areas engaged by an fMRI emotion processing and regulation paradigm in synergy with Project 1 (P1), and these changes will partly explain exercise-induced improvements in positive and negative affect measured in daily life by EMA and by conventional self-report instruments in synergy with Project 2 (P2).

#### 1.1 BACKGROUND AND RATIONALE:

PA is one of the most critical behaviors to reduce risk for many chronic diseases, including some cancers (e.g., breast cancer), cardiometabolic conditions (e.g., Type II diabetes), and atherosclerotic cardiovascular disease (CVD)[4-6]. Moreover, PA has beneficial effects on a range of proximal mediators and markers of chronic disease risk—especially CVD risk. These include blood pressure (BP), cardiac autonomic control, inflammation, glucose regulation, adiposity, and lipid levels. In patient samples, clinical trials show that PA accelerates recovery from myocardial infarction[8], coronary bypass surgery and congestive heart failure[9], improves quality-of-life, and reduces risk for new CVD events and all-cause mortality[10]. Further, cardiac patients who improve their fitness show a 20% reduced mortality rate over an 11-year follow-up period[11]. Based on an expansive body of evidence, the American Heart Association (AHA) has identified CRF as an important 'vital sign', and it has recommended more frequent assessments and management of fitness by health professionals[12,13]. Finally, physical *inactivity* is estimated to account for >350,000 deaths per year in the USA alone, with an additional 3.2 million annual deaths globally[14]. In view of clear scientific evidence on PA and health, many international organizations have nearly identical recommendations for people to engage in PA. The most recent were established in 2018, advising >150 min of moderate intensity exercise, or at least 75 min of vigorous exercise, per week to improve cardiovascular health and to reduce risks for premature mortality[2]. Less widely appreciated by health care professionals, the public, and policy makers are the broad benefits of PA on the brain. These benefits translate into brain-based improvements in cognitive, affective, stress-buffering, and self-regulatory behaviors [5,7]. Indeed, substantial human and animal evidence shows that the brain exhibits appreciable structural and functional plasticity (i.e., change) from PA. As a result of this evidence, many organizations that influence public policy have developed campaigns to educate the public and health care professionals on the brain benefits of PA. For example, the NHLBI and AHA note mood improvements and reductions in psychological distress as established benefits of engaging in PA [15]. In support of these health campaigns, Barnes & Yaffe[19] further reported that >1.1 million Alzheimer's cases in the USA are potentially attributable to physical inactivity. Noteworthy here is that Alzheimer's and other forms of dementia are highly co-morbid with CVD, and they are often predicted by CVD risk factors that are affected by PA (e.g., hypertension)[16-21]. Moreover, exercise reduces aspects of negative affect and depressive symptoms that associate with CVD risk[22-24]. Finally, there is preliminary evidence to suggest stress-buffering effects of PA and cardiorespiratory fitness at the level of physiology and behavior, which may define additional pathways to reduce CVD risk. *What is not known, however, is whether the beneficial effects of exercise on the brain extend beyond emergent cognitive, affective, and stress-buffering processes to also include beneficial effects on physiological and systemic mediators of CVD risk that are regulated by the brain. If so, such effects may help to more precisely define and refine novel and brain-based targets for intervention and prevention efforts.*

#### 1.2 INNOVATION:

We have assembled a productive team with a history of collaboration in exercise interventions. eBACH questions are unique in the field, potentially transformative, and relevant to both scientists and health professionals.

- **Body-to-brain pathways linking physiology and exercise.** For the first time, we will test whether exercise-induced changes in peripheral markers of CVD risk explain changes in brain plasticity.
- **Brain-to-body pathways linking exercise and physiology.** We will test the novel hypothesis that the effects of exercise on brain circuits supporting visceral control account for changes in autonomic, hemodynamic, and neuroendocrine mediators of CVD risk.
- **Dense repeated measurement of exercise and neural-CVD risk associations.** For the first time in a randomized exercise intervention, we will collect 3 measurement points of high-resolution neuroimaging data and peripheral mediators and markers of CVD risks. This allows us to more accurately model trajectories of change for both neuroimaging and CVD risk related measures, enabling us to move beyond traditional correlation, observational, and briefer wave assessments of these associations.
- **Statistical modeling of exercise and CVD risk.** We have the expertise to apply cutting-edge machine learning approaches to neuroimaging, behavioral, and peripheral biomarkers to develop a more comprehensive and realistic model of the associations between exercise and its associations with peripheral physiology, risk factors for CVD, and brain plasticity.
- **Neurobiology of exercise and emotion and stress linked to CVD risk.** We will examine how exercise alters stress physiology and affective CVD risk correlates via changes to brain circuits.

#### 1.3 PRELIMINARY DATA:

1. **Peripheral mediators and markers of CVD risk and PA**: In the AHAB2 registry, we tested associations between markers and mediators of CVD risk and PA after control for age and sex. In AHAB2, energy expenditure (assessed over ~5 days by actigraphy) was negatively correlated with mean carotid intima-media thickness ($p = 0.017$, $N = 437$) and an aggregate metabolic risk score ($p < 0.001$, $N = 414$). Also, energy expenditure was positively correlated with an index of cardiac vagal control derived from high-frequency heart rate variability (HF-HRV) and root mean square of successive differences (RMSSD) measured during paced and unpaced breathing epochs ($p < 0.001$, $N = 432$). Moreover, greater energy expenditure correlated with lower interleukin 6 (IL-6) and C-reactive protein (CRP) (ps < 0.001, Ns = 453 and 454). The cardiac vagal control index also negatively correlated with IL-6 and CRP (p's < 0.005, Ns = 448 and 449). In line with our aims, energy expenditure was positively correlated with bilateral hippocampal volume after additional control for intra-cranial volume (ICV) ($p = 0.024$, $N = 425$). Finally, the cardiac vagal control index was positively correlated with volumes of the ACC, amygdala, and hippocampus (ps < 0.05). *Implications for this project*: These results show associations between PA and peripheral markers and mediators of CVD risk, and that these associations may be linked to markers of brain plasticity and health. What is unknown, and what is tested here, is whether such cross-sectional associations between CVD risk markers, PA, and brain health are causally linked in the context of an experimental manipulation of PA.

2. **PA, stress, and affect**: In brief, we replicate several observations from the literature on associations between PA and fitness with affect, and we provide the first

evidence that PA relates to blunted stressor-evoked BP reactivity in daily life. We also report the first associations to our knowledge on PA and individual differences in neural activity during the processing of negative emotional stimuli, as well as during the regulation of negative emotions. In aggregate, these initial findings support the integration of the fMRI stressor and emotion task batteries, as well as ambulatory (ABP) and ecological monetary assessment (EMA) methods.

3. **Exercise and brain health**: Our research group has published >170 papers on the effects of exercise and fitness on brain morphology, white matter integrity, resting state connectivity, and task-evoked functional MRI activity. But, none of these have specifically addressed the questions at issue here. Evidence from our work and others reliably demonstrates that PA and CRF are related to several metrics of brain health across the lifespan in clinical and healthy samples. For example, we found in children (N=35/group) that exercise is associated with changes in task-evoked fMRI[29], cerebral blood flow in the hippocampus[30], and increased hippocampal volume[31]. Similar effects of exercise are seen in young adulthood and mid-life. For example, we found in a randomized trial of PA (N=125) in adults between 18-50 yrs that 12-months of PA increased cerebral blood flow in the ACC, insula, and PFC. Increases in hippocampal and PFC volume is also seen in young adults after 6-months of aerobic exercise[32]. Similar positive effects of PA have been reported in mid-life adults with T1 diabetes[33], in multiple sclerosis patients[34], and in women with breast cancer[35]. Positive effects of PA on brain health are often observed in late-life. For example, in 120 older adults (60-80 yrs; N=60/group), moderate intensity exercise increased hippocampal volume. Exercise also increased volume of the PFC and ACC in 59 (N=~28/group) older adults[26]. In 879 older adults, greater energy expenditure was associated with greater volume of the hippocampus, ACC, and PFC[36]. In addition to brain morphology, changes in CRF resulting from an exercise intervention were correlated with increased white matter integrity in PFC and temporal lobes in 70 older adults[28] and also in two larger samples (N=113; N=154). Our results also extend to resting-state connectivity[27,37,38] and task-evoked activation[39]. For example, in 65 older adults exercise increased functional connectivity[27]. Overall, this rapidly growing literature indicates that PA, CRF, and aerobic exercise in particular, is effective for improving several metrics of brain health in visceral control circuits and that these effects occur across the lifespan in diverse samples.

#### 1.4 STATISTICAL ANALYSIS:

**1.4.1. POWER:** Our proposed intervention is much larger and much longer than other similar well-controlled randomized PA interventions, especially those that include neuroimaging. Prior studies are typically <6 months in duration with N<25/group. Thus, our intervention with 3 measurement points (0, 6, 12 months) with N=150 (75/group) far exceeds other comparable studies in this area. Well-controlled RCTs typically have smaller sample sizes than traditional observational studies because of the increased power obtained through the within-subject analytical approach. Indeed, the number of assessments (450) is similar to that of Projects 1 and 2. Here we describe the statistical basis for our proposed design using an estimated sample size after 20% attrition (N=60/group). Brain volumetrics: Our prior 6-12 month PA interventions in children, young adults, and older adults have used between 20[26] and 60[40] subjects per group with moderate-sized effects (Cohen's d) ranging between 0.30 and 0.50. Cross-sectional research has supported these numbers with studies reporting significant effects of PA on brain volume with only 32 subjects[41] and slightly larger effect sizes between 0.50 – 0.60. Even if we estimate a rather conservative effect size of 0.30, 60 participants per group is sufficient to detect differences in brain volume at 80% power. Diffusion imaging: Changes in white matter have been detected in PA interventions in as little as 6-months with only 50 subjects[26]. In a 12-month exercise intervention, we found increased FA with only 35 subjects in each group with an estimated effect size of 0.70[28]. Fewer results have been reported in mid-life, but in children, effects of PA on white matter have been reported with N=25/group. Based on an estimated effect size of 0.75, we should have 90% power with 60 participants per group to test our main hypotheses as well as enough power for exploratory analyses. Connectivity: With only 65 people, we have shown that a 12-month exercise intervention is effective at enhancing functional connectivity in prefrontal-inhibitory networks[27]. The difference between the groups for PFC connectivity was .15 with a standard deviation of .10 suggesting an effect size of 1.5 (Cohen's d). In young adults, we report similar effects with N=50 with increased functional connectivity between the anterior hippocampus and ACC. Therefore, similar to the effect sizes and power for the other neuroimaging measures, 60 participants per group should be sufficient for detecting changes as a function of the intervention with 95% power. Stress reactivity and affect: In preliminary data from the AHAB2 cohort, greater PA was associated with reduced daily life stress reactivity measured by strain (r=.23) and conflict (r=.25). In a 12-week exercise intervention in young adults with major depression scores on the Montgomery Asberg Depression Rating Scale significantly decreased from 21.8 to 4.25, and the magnitude of this decrease was correlated with increases in fitness (r=-.39). Based on these effect size data, 60 participants per group would provide us with >90% power to detect an effect on stress and affect measures. Thus, we estimate adequate power to test Aim 2. CVD markers: Prior randomized exercise interventions examining some of the same CVD risk markers proposed here (e.g., baroreflex sensitivity [BRS]) are typically between 4-52 weeks in duration with N= ~10 and 150[31]. Despite this heterogeneity, meta-analyses of exercise interventions show moderate-to-large effects on many of these outcomes (Cohen's d=.60). For example, a meta-analysis of 30 exercise interventions on multiple CVD risk markers reported an average 18 week duration with ~50 subjects in each study (~25/group)[25]. In this meta-analysis, CRF increased by 3.04 mL/kg/min with a Cohen's d of 0.58. We expect larger effect sizes here because durations of ~1-year demonstrate larger changes in these metrics[42]. Thus, based on prior studies and meta-analyses, we are sufficiently powered with N=150 to test for exercise-induced changes in CVD markers. Mediation: Using established empirical estimates of sample sizes needed for .80 power in bias-corrected bootstrapping analyses with moderate sized effects in both alpha and beta, it is estimated that 115 participants are needed[43]. Thus, based on the above effect sizes for both brain and CVD markers, we are sufficiently powered to test our hypotheses using structural equation modeling even after attrition. In sum, our planned sample size of 150 (N=75 per group) will be sufficient for allowing us to test our primary hypotheses.

**1.4.2. PRELIMINARY STEPS**: Prior to hypothesis testing, all data will be examined to determine: 1) frequency distributions for missing data and out-of-range values; 2) normality and internal consistency of subscales; and, 3) association between variables that may be highly correlated to guard against multicolinearity, which would inflate standard errors and make estimation unstable. We will also examine the distribution of factors between groups to assess whether covariate-adjustment is required. *Missing data*: All outcomes will be tested using an intent-to-treat (ITT) framework. Sensitivity will be explored using adherence to the intervention (e.g., per protocol). We will report reasons for dropouts and explore missing data mechanisms. To examine whether missingness is unrelated to other observed and unobserved measurements (missing completely at random, MCAR) or to the observed measurements only (missing at random, MAR), testing for completely random dropouts will be carried out. Additional sensitivity analyses test for informative dropouts. We will apply pattern-mixture models[44] by stratifying our data by dropout patterns and fitting separate regression models to strata.

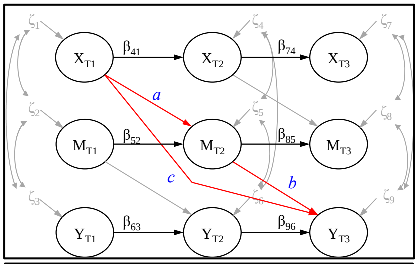

**Figure 1**. Diagram of exemplar full longitudinal mediational model. Paths involved in estimating longitudinal stability and residualized change are in black, paths involved in mediation are in red, and non-essential parameters are grayed out for legibility.

**1.4.3. PRIMARY OUTCOMES**: Before testing the specific aims related to mediation we will test whether the intervention modifies proximal endpoints that will serve as the primary outcomes of the study. Thus, the primary endpoints will be 12-month change scores resulting from three different measurements: (1) hippocampal volume, (2) mean systolic blood pressure reactivity averaged from Stroop and Multi-Source Interference Task (MSIT) stressors during fMRI, and (3) heart rate variability. More specifically, we are predicting Time x Group interaction terms such that the exercise arm would change more than the control arm for these measures. Changes in scores of other measures including EMA-derived measures of daily life positive and negative affect, BRS, pulse wave velocity [PWV], vasodilation, CRF, resting state brain connectivity, and cortical thickness will be considered secondary and tertiary outcomes. Further, results from the structural equation modeling will be considered secondary outcomes, but will be testing the specific aims of the project.

We propose to test our primary hypotheses using a general linear mixed model (GLMM) approach because we will have three assessments for each of our measures over the course of the trial (baseline, 6-months, 12-months). The

GLMM model will include both random intercept and random slope for individual participants and a treatment-by-time interaction as a fixed effect. This technique will allow us to model the changes in the scores as a function of both time and group while also including other potentially confounding variables in the model. GLMM modeling of repeated assessments is a popular approach in clinical and longitudinal studies because it allows for random changes in measurement error and takes into account both within and between subject correlations and estimates the average growth curve across subjects and subject-specific growth curves.

**1.4.3 <u>MEDIATION</u>:** Aim 1 has two hypotheses, (a) to test whether the peripheral effects of exercise mediate brain effects, and (b) whether the effects of exercise on the brain mediate the beneficial effects of exercise on peripheral mediators of CVD risk. We propose to test these hypotheses using longitudinal mediation models in a structural equation modeling (SEM) framework. *<u>Aim 1A:</u>* In this model, the exercise groups (X) are proposed to influence visceral control circuits (Y) via changes in peripheral pathways (M) including insulin resistance, oxidative stress, endothelial dysfunction, and CRF. We will first examine whether brain or these peripheral CVD pathways can be summarized as latent variables to control for measurement error. When using latent variables, the modeling will proceed by first establishing well-fitting measurement models, and testing for measurement invariance across waves. To test the hypothesis proposed in Aim 1A, analyses will be conducted both on a voxel-wise basis (e.g., using the BRAVO Toolbox (https://sites.google.com/site/bravotoolbox/)) or using extracted data from SPM, FSL, or FreeSurfer, depending on the imaging modality and analysis. In the latter case, we will use a theory-driven approach by selecting values from anatomical or functionally-defined regions-of-interest (ROI) on visceral control regions. Each imaging modality will be analyzed in separate models unless we have a theoretical or empirical basis for including multiple imaging metrics in a single model. We will use standard cluster-based and FDR corrections for multiple comparisons for all voxel-wise analyses and the Brain Connectivity Toolbox for graph theory metrics (i.e., segregation). Although directional hypotheses have been posited for our mediation analyses, a more conservative significance will be set at .01 to limit inflation of type 1 error. Global and incremental model fit will be verified using multiple fit-indices (chi- square, RMSEA, CFI, TLI, SRMR). With three waves of data, we will use full longitudinal mediation to test our hypotheses. Full longitudinal mediation allows us to test the effect of initial standing on the hypothesized predictor (X – Exercise group) at T1 on change in the mediator (M – peripheral CVD markers) from T1 to T2, controlling for initial standing on M, and the effect of the mediator (M– peripheral CVD measures) at T2 on the change in the outcome (Y – change in brain measure) from T2 to T3, controlling for initial value of the outcome. Mediation will be tested by calculating the indirect effect of paths that are consistent with longitudinal mediation (i.e., paths *a* and *b* in **Fig. 1**). Here the estimate *a**b* provides a measure of the longitudinal mediated effect because it represents the effect of X at T1 on residualized change in M (path *a*), and M at T2 on residualized change in Y (path *b*). Significance of the mediation (i.e., indirect) effect will be examined using techniques that account for non-normality in the parameter distribution, namely bias corrected bootstrapping-based approaches. A benefit of full longitudinal mediation models is that the direction of the paths can be reversed (Y->M->X) to test for the opposite causal direction, and bidirectional effects can similarly be tested. We will test for these possibilities by comparing models with paths reversed and allowing bi-directional effects. Finally, we aim to explore the role of other potential mediators (i.e., sleep) and moderators (e.g., gender) on the direction and strength of associations. *Aim 1B*: We will proceed with similar approaches described above for Aim 1A including using a voxel-wise approach and a theory-driven approach by selecting anatomical or functionally-defined ROIs on visceral control circuits or the white matter pathways that connect them. Importantly, there may be regional dissociations between the brain regions identified in Aim 1A and those identified in Aim 1B. Using voxel-wise analyses, we will be able to adjudicate the regional specificity or overlap between the regions identified in Aim 1A with those of Aim 1B. Using these procedures, we will also be able to determine the specificity of particular effects (e.g., regions associated with BRS but not with HRV) or differences across imaging metrics (e.g., differences of effect between functional connectivity, cortical thickness, white matter). With three waves of data, we will use full longitudinal mediation to test our hypotheses that X (Exercise group) influences BRS, HRV, and glucocorticoid control (Y) via its effects on brain plasticity on visceral control circuits (M). The description of the SEM modeling approach for Aim 1A will be applied to test mediation for Aim 1B. *To test Aim 2A & B*: The analytical plans for testing Aim 2A and 2B along the lines depicted in the heuristic model of Figure 1 will proceed like that described above for Aim 1A and B. That is, we will test how the effect of the hypothesized predictor (X – Exercise group) influences the change in the mediator (M – visceral control circuits), on stress reactivity and affect metrics that are measured in Projects 1-2 (e.g., EMA and ABP measures identical to those used in NOAH). Similar to Aim 1, we will be able to model opposite causal directions and bidirectional effects by reversing the paths in the model and will be able to determine specificity of particular effects (e.g., regions associated with emotion regulation, but not daily life stress reactivity) or differences across imaging metrics (e.g., differences of effect between functional connectivity, cortical thickness, white matter). As above, we will also test other potential mediators (i.e., sleep) and moderators (e.g., gender) on the direction and strength of associations.

#### 1.5 PROCEDURES:

Before the collection of baseline data, all staff will have had ethics training and appropriate certification of research training modules.

##### 1.5.1 SCREENING:

During the UCSR telephone screening interview, numerous medical history questions will be asked to verify that the potential participant meets inclusion criteria as no medical record retrieval is being used. Information about safety to exercise will be obtained over the phone (i.e., history of falls) to ensure that exercise will not be dangerous for the participant. Once deemed eligible, this participant's contact information will be sent to an eBACH staff member who will conduct a secondary screen specifically for physical activity and MRI safety eligibility. Information about the participant's current exercise status will be collected and participants that engage in more than 75 minutes of structured exercise per week will be considered ineligible. Information regarding metal in the body for MRI safety will obtained, including information about any metallic objects in their body. If a participant is deemed unsafe to exercise based on the physical activity readiness questionnaire, the participant will be required to have prior approval by their primary care physician before being admitted into the exercise program.

##### 1.5.2 INCLUSION AND EXCLUSION CRITERIA:

**Inclusion Criteria:**

- **Age** – individuals aged 28-56 years old when signing the consent form.
- **Gender & Ethnicity** – men and women are both eligible to participate. We expect the study population to be approximately 50% women. All ethnic groups are eligible for the study. We expect the study population to be approximately 30% minorities.
- **Ambulation** – study participants must be able to walk without pain or use of an assisted walking device. This will be determined during the initial phone screen. Ambulation will also be assessed in person as the potential subject must be able to complete the $VO_{2\,Max}$ test on a treadmill.
- **Physical Activity and Fitness Level** – all participants must report engaging in less than 75 minutes per week of exercise and have a $VO_{2max}$ percentile less than 75 based on the ACSM Guidelines for Exercise Testing (10$^{th}$ edition).
- **Residency -** Since the intervention is 12 months in duration, all potential participants must reside in the Pittsburgh area and plan on residing in the area for at least one year after randomization. It is recommended that the study coordinator during the second screen assess potential participants' distance from the exercising sites since the study requires 2 visits per week. The further a participant is from an intervention site, the higher the probability is for the person to be unable to adhere to the protocol.
- **Reliable transportation:** Participants must have reliable transportation to complete the outcome measures and the intervention requirements.
- **Willingness to be randomized -** To be eligible, all potential study participants must be willing to be randomized to either of the intervention arms. Individuals that insist on participating only if they can choose which arm they belong to should not be enrolled.

**Exclusion Criteria:**

- Medication Use:
  A. Reported use of the following medications on one or more occasions in the past 14 days constitutes grounds for exclusion:
    1. Antihypertensive or cardiac medications (diuretics, beta blockers, calcium channel blockers, ACE inhibitor/ARB, cardiac glycosides, central sympatholytic HTN drugs, anti-arrhythmic drugs, vasodilator drugs, other cardiac drugs).
    2. Anticonvulsant medications
    3. Antiparkinson medications
    4. Protease inhibitors or other Anti-HIV medications
    5. Medications for the treatment of mania
    6. All other centrally active or psychotropic medications excluding anti-anxiety, antidepressant, or antipsychotic/tranquilizer medications.
    7. Insulin
    8. Chemotherapy
    9. Immunosuppressants and related biological agents (Imuran, methotrexate, and cyclophosphamide)
    10. Prescription weight loss medications and ephedrine OTC

  B. Reported use of the following medications on a “regular” basis is grounds for exclusion. For this purpose, “regular use” is defined as reporting that the prescribed medication was taken 7 or more days in the past 14 days. Individuals who take these medications, but less frequently than 7 out of the past 14 days are not excluded:
    1. Anti-anxiety medications (Ativan, valium, Xanax)
    2. Sleep medications (e.g., trazadone at bedtime for sleep)
    3. Asthma oral medications
    4. Asthma/allergy inhalants (alupent, albuterol)
    5. Anti-depressant medications
    6. Antipsychotic/tranquilizer medications
    7. Glucocorticoids (oral; prednisone, cortisol)
    8. Medical marijuana

  C. Reported use of more than 2 non-insulin medications for diabetes on a “regular” basis is grounds for exclusion. For this purpose, reported use of combination medications, involving two or more non-insulin medications for diabetes in a single pill, is counted as 2 separate medications and would be grounds for exclusion.

  D. A person who reports that he or she was once on a disallowed medication, but has discontinued this medication for at least a month or longer and is otherwise eligible, is allowed to participate in the study.
- Substance use exclusions:
  A. Anyone reporting 35 or more alcoholic drinks in the last 7 days is excluded.

B. Anyone reporting consumption of 6 or more alcoholic drinks on 3 or more occasions in the past 7 days is excluded.
C. Anyone reporting a score of 8 or more on the 10-item AUDIT scale (indicating harmful or hazardous use of alcohol) is excluded.
D. Anyone reporting use of illicit drugs on 7 or more days in the past 2 weeks is excluded.

- Medical conditions that are disallowed are:
  A. Self-reported prior heart attack, stroke, bypass surgery, angioplasty, congestive heart failure, arrhythmia (cardiac rhythm problems).
  B. Hypertension (SBP/DBP $\geq$ 160/and/or $\geq$100 mmHg).
  C. Cancer (treatment in last 12 months, allowance for non-melanoma skin cancer) - potential participants who have received treatment in the last 12 months for cancer, including radiation or chemotherapy will be excluded from the study.
  D. Liver disease - any person who has had hepatitis B or C, liver failure or Cirrhosis will be excluded from the study.
  E. Kidney disease - potential participants who indicated they have chronic kidney failure, have undergone dialysis, or have had a kidney transplant will not be eligible to participate.
  F. Type I diabetes or insulin dependent Type II diabetes - Type 2 diabetics if they are taking insulin or 3 or more diabetes medications. Single injections that contain 2+ drugs count as 2 medications.
  G. Self-reported history of a major neurological disorder or brain injury resulting in ongoing symptoms or cognitive impairment (e.g., multiple sclerosis, cerebral palsy, major head injury)
  H. Self-reported chronic psychotic illness (schizophrenia, bipolar disorder).
  I. Lung disease requiring drug treatment (asthma or allergy inhalers are <u>not</u> exclusionary unless they are used on a "regular basis")
  J. Weight loss surgery within the past 5 years.
- Other exclusionary criteria:
  A. Current pregnancy or plans to become pregnant over the next year – any woman who is pregnant via a pregnancy test at the baseline visit will be excluded from the study. A pregnancy test will also be given at the MRI visit and this will be repeated at 6-months and 12-months.
  B. Those who make regular use of an assistive walking device are excluded.
  C. Those who are not fluent in English are excluded (defined as having spoken and read English every day for <10 years).
  D. Those with a visual impairment that would prevent them from reading printed text or text on a computer screen, iPad, or other electronic device are excluded.

E. Those with color blindness, who may not be able to distinguish colors on some of the tasks used in this study, are excluded.
F. Those who have claustrophobia (fear of enclosed or confined spaces) are excluded.
G. Those who have medical devices, implants or other metal objects in or on the body that cannot be removed and are incompatible with use of fMRI (for example, tattooed eyeliner) are excluded.
H. Those who are unable to fit into an MRI scanner are excluded.
I. Those who report working the night shift on a frequent basis (half or more of the hours worked in a full workday are between midnight and 8 am, and this has occurred more than 12 times during the past year) are excluded.
J. Those without reliable access to a telephone throughout the day (home, work, or cell phone) are excluded.
K. Those who are otherwise unable to meet the requirements of the study (for example, persons whose employment or personal situation will not permit momentary interruptions required for electronic diary and ambulatory data collection) are excluded.
L. Those unwilling to be randomized to one of the two groups will be excluded.
M. Individuals that self-report engaging in 75 minutes or more of structured physical activity per week will be excluded. This will be determined using a combination of questionnaire and interrogation by trained exercise staff.
N. Those that have cardiorespiratory fitness levels that are above the 75%ile based on ACSM criteria will be excluded.

##### 1.5.3 <u>BASELINE TESTING:</u>

There are multiple outcome measurement sessions at baseline. There is an 8-week maximum window to complete these measures. The 8-week window starts the day the informed consent is signed at session 1. Any measures or sessions collected outside of this window would not determine eligibility, but would be classified as a protocol deviation. Further, the order of these sessions and measures might be re-ordered based on availability and timing of available slots.

**<u>Session 1: ELIGIBILITY AND FITNESS TESTING</u>**

Before the session, participants will receive a reminder phone call and will be instructed to refrain from exercise or drinking alcohol after 9:00 pm the night before, no over-the-counter medications 12 hours prior to the appointment, no smoking, using nicotine, or consuming caffeine 3 hours before the visit. Informed consent will be obtained during the first session (~ 3 hours). Participants will have their blood pressure taken (clinical read) to re-verify eligibility. Measurement of the participants height and weight will be taken using a validated stadiometer. Participants will be asked to bring their medications in a bag for verification. Once the participant is deemed eligible, they will fill out several questionnaires before undergoing a CRF test. Participants will complete a maximal $VO_2$ test to assess CRF following a Modified Balke Protocol with an agreed upon speed between the participant and lead exercise physiologist. The session will first start by a warm-up session of 5 minutes followed by a blood-pressure reading. Only participants with safe blood pressure readings (>220/110) will be allowed to continue. The participant will walk on a motor-driven treadmill with constant speed and increments of the incline. The intensity is increased in two-minute stages. We will collect all of the exhaled air by having the individual wear a fitted facemask that collects the exhaled air which then travels through a tube to the analyzer. This assessment usually takes about 15 - 30 minutes to complete depending on the starting fitness level of the participant. These procedures are recommended by the American College of Sports Medicine for graded exercise testing. During the exercise test, heart rate is continuously monitored via ECG along with blood pressure readings and Rating of Perceived Exhaustion (RPE) every two minutes. When the participant reaches the endpoint goal of the exercise test (symptom limitation and/or volitional exhaustion), the facemask will be removed and they will undergo a four-minute active cool-down in which they will walk at a slower rate with zero incline grade. After these four minutes the participant will be helped off of the treadmill and will undergo a passive cool-down session in which they are seated in a chair. The participant's heart rate and BP will be continuously monitored through the cool-down period every two minutes and the participant will not be allowed to leave until the vital signs return to baseline patterns (within a value of 20 mmHg of systolic and diastolic and 20 beats per minute of heart rate relative to their baseline reading).

Each maximal exercise test will be administered by a trained exercise physiologist and exercise technician. The exercise technician should be the staff member who records the required values of each stage and assists with test administration. The exercise physiologist should be the staff that leads the exercise test and provides verbal cues to the participant. The exercise testing will be used to measure changes in fitness levels after 6-months and 12-months. The room should be set at a comfortable temperature (recommended between 60 – 70 degrees Fahrenheit), have necessary fans in place, and have access to water for the participants. The exercise physiologist will also be responsible for examining the ECG read-out and anyone displaying any questionable patterns will have the ECG read by a consulting cardiologist for the team for confirming safety or need for additional follow-up with the participants PCP or cardiologist. Continued eligibility for the study will be dependent upon this consultation and follow-up.

Also at this visit, participants will be fitted with a commercially available physical activity monitoring device (Actigraph Link) around the non-dominant hand wrist that will record objective physical activity data over the duration of approximately one week. The participants will be provided with detailed instructions regarding wearing of the device as well as the option to remove the device if it becomes problematic. Participants will complete a series of questionnaires at this visit.

Once the $VO_{2max}$ session is complete, participants will be given the NCI diet history questionnaire, via a link and study code that is unique to each participant. Participants will have the option to complete the questionnaire online at their home or via a paper based method to be returned to the eBACH staff.

**Session 2: EMA INFORMATION**
This visit will take approximately three hours and the entire session will occur at Sennott Square in Oakland. The visit entails a clinic blood pressure assessment, and therefore participants will follow the same pre-visit restrictions as visit 1 (no exercise or drinking alcohol after 9:00 pm the night before, no over-the-counter medications 12 hours prior, no food or drink (except water), no smoking, using nicotine, or consuming caffeine 3 hours before the visit). Participants will be notified of these restrictions by a phone call made by a research assistant at least one day prior to the visit. This call and all calls to the participant during the daily monitoring phase will be made from the study cell phone.

Additionally, participants will complete several questionnaires assessing their physical and emotional well-being.

At this visit participants will also receive training on how to use several ambulatory monitoring devices that they will carry with them during the 10-day daily monitoring phase of the study. These devices will measure physical activity and sleep (actigraphy wristband), ambulatory blood pressure (portable blood pressure device, belt, and cuff), and daily psychosocial stressors (Diary of Ambulatory States [DABS] completed on a smart phone). Participants will receive detailed instructions on how to use these monitoring devices, including an option for removal if any of them present a problem. Following the 10-day monitoring period, we will collect the devices, and after we download the data we will clean and sanitize them so that they can be used by other participants in the study. While participants wear the devices they will not be able to view any data.

After the training session, participants will leave the lab practice using the monitoring equipment for the rest of the day and for the morning of the following day. Around late morning, a research assistant will call participants for their 15-minute "Practice phone call," in which they will be given assistance with any problems or questions that may have come up during the practice period.

Following the practice phone call, participants will be instructed to begin the 10-day monitoring phase. This phase consists of four "full" monitoring days (using all devices and completing all interviews) and six "partial" monitoring days (using only the actigraphy device and the electronic diary to complete a morning interview and an evening interview).

On the "full" days, participants will have their blood pressure taken every hour they are awake. Following each blood pressure reading, they will complete a 2-5 minute questionnaire (DABS) about their thoughts and feelings using the electronic diary. If the participant uses nicotine, they will be trained to activate the electronic diary every time they use a nicotine product. Participants will wear the actigraphy wristwatch while they are awake and asleep. Before participants go to bed, they will be prompted to turn off the blood pressure device and set a wake-up alarm on the electronic diary for the following morning. Once they wake up from this alarm the following day, they will turn back on the ambulatory blood pressure monitor and put it on within 45 minutes. These 45 minutes allow for grooming, showering, and other morning routine activities. On two of the "full" monitoring days, participants will also wear the blood pressure monitor overnight, however they will not need to complete the hourly electronic diary questionnaires overnight.

On the "partial" monitoring days, participants will not wear the blood pressure monitor or complete the hourly electronic diary interviews. The actigraphy wristwatch is worn on both "full" and "partial" monitoring days of the study (including every night of sleep during the monitoring phase).

Several additional phone calls will be scheduled for the first or second day of the monitoring phase in order for a research assistant to answer any of the participants' questions and asses any difficulties they may be having. Similarly, another phone call will be made on the third or fourth day of the monitoring phase in order to confirm that the equipment is still working correctly and to review instructions with the participant.

On two separate non-"full" monitoring days during the daily monitoring period, participants will be instructed by a research assistant to complete the Automated Self-Administered Recall System (ASA24) outside the lab.

**Session 3: CARDIOVASCULAR HEALTH**
This visit will take place at Old Engineering Hall, will last (~3 hours), and will be conducted by a trained researcher. It involves returning the EMA equipment to research staff for data retrieval, assessment of pulse wave velocity and endothelial function, and additional questionnaires.

EMA Device return:
Participants will arrive to the visit wearing their devices. They will take off their devices and a research assistant will review the data that was collected over the 10-day period. If it is determined that the participant did not complete the expected number of interviews and blood pressure readings, they will be asked to complete extra day(s) of monitoring to provide complete assessments.

Participants will be asked about their compliance with pre-visit instructions (same as Visit 1 plus an additional instruction to wear loose clothing that allows research staff to attach heart-activity measuring devices without disrobing). Similar to the previous visits, these instructions will be given during a phone call made by a research assistant the day prior to the visit. While the research staff is reviewing the participants' daily monitoring data, participants will complete a number of psychosocial questionnaires.

Cardiovascular Assessments:
Following the return of the EMA devices, three separate (non-invasive) protocols will be administered to assess aspects of the participant's cardiovascular health/function:

*Autonomic Function and Arterial Stiffness:* Self-adhesive sensors will be applied to the participant (on the back of the neck, upper chest, shoulders, ribs, and calf area below the knee) to record their electrocardiogram and monitor changes in thoracic/calf impedance due to blood flow (dual-impedance cardiography). Continuous beat-by-beat BP will also be monitored using cuffs placed around the participant's non-dominant arm and the index/middle fingers of the opposite hand. Recordings will be taken for 5 minutes while the participant is at rest and breathing normally, and also during a paced-breathing protocol (i.e., 11 breaths per minute). From these recordings we will gather key measures of HRV, BRS, and pulse wave velocity (PWV).

*Endothelial Health:* Venous occlusion plethysmography (VOP) will be used to assess the participants' endothelial function. First, participants are comfortably seated and outfitted with the necessary equipment, including: an occlusion cuff for both their wrist and arm, and a strain gauge wrapped around their forearm. During a standard sequence of cuff inflations/deflations, the participants' forearm blood flow is measured via changes in forearm circumference (as indexed by the gauge). Following "baseline" readings, the arm cuff is inflated to 40 mmHg above systolic pressure for 5 minutes until it is released and "max" readings re-commence. This post-occlusion state is referred to as reactive hyperemia and provides a reliable index of endothelium dependent vasodilation. The participant's max flow readings are compared to their baseline readings to derive a percentage increase, with greater values associated with better health.

*Pulmonary Function:* Participant's lung function will be measured using spirometry. While standing, individuals are instructed to inhale/exhale into a mouthpiece under normal and forced/maximal conditions. From these tests a participant's forced volume capacity and forced expiratory volume can be determined.

*Blood Draw:*
Lastly, a trained phlebotomist under the supervision of Dr. Anna Marsland (Co-I) will conduct a blood draw for inflammatory and metabolic measures and a hair collection sample to assess hair cortisol levels.

Site specific laboratory safety policies when handling human blood samples will be followed including the use of Biological Safety Cabinets and Personal

Protective Equipment/PPE: gloves, lab coat/gown, glasses, and face mask at all times when working with specimens in the lab.

Participants will be asked to abide by the following fasting restrictions prior to the session: nothing to eat or drink (including alcohol and caffeine) except water from 9pm the night before; no exercise for 12 hours prior to the session; no nicotine/smoking 3 hours prior to the session; no cold/allergy or headache medication (aspirin, ibuprofen or other NSAID) for 12 hours prior to the session. A researcher will call the participant the night before the appointment and ask if they have taken antibiotics in the past 2 weeks, or if they have a current infection or cold symptoms (see attached Blood Draw Eligibility Questionnaire). If so, they will be rescheduled. Participants will also be rescheduled if they have had anything to eat or drink except water since 9pm the night before their appointment or report acute illness symptoms.

Blood will be delivered to the Behavioral Immunology Laboratory (605 OEH) of Dr. Anna Marsland within 30 minutes of the time of the blood draw. Blood will also be delivered to UPMC Central Labs for glycemic and lipid assays.

*Hair strands:* ~ 50 mg; 100-150 strands will also be collected with fine scissors as close as possible to the scalp from the upper part of the back of the head so that it is not noticeable. The samples will be retained by the Behavioral Immunology Lab and tested for cortisol levels.

**Session 4: MRI**

The fourth session will be an MRI Scan. It is anticipated that the complete session will take ~3 hours to complete, with less than 60 minutes in the scanner. Participants will be scheduled for the MRI scan and standard sequences will be collected that will consist of images of brain structure and function.

This session will take place at the BRain Imaging Data Generation & Education (BRIDGE) MRI Center located at the Mellon Institute and will be conducted by a trained researcher. Once at the center, participants will have their seated BP and HR taken, and they will receive training for the MRI protocol that will be administered. An additional screening for MRI eligibility will take place as a safety precaution, in addition to a pregnancy test (if necessary). Participants will also be asked to remove any piercings and completely empty their pockets of all contents. They will be given ear protection against the sound of the MRI machine and asked if they need to use the restroom before beginning.

Subjects will also be equipped with physiological monitoring (EKG sensors, a respiration belt, and a BP cuff) devices before being put into the scanner. The total time in the scanner will not exceed 60 minutes.

MRI Protocol:

Structural MRI sequences: Several structural sequences will be collected that will provide detailed information about brain morphology (T1 MPRAGE sequence) and hippocampal morphology (high-resolution hippocampal sequence). During these scans, participants will be asked to lay still and relax quietly.

Functional MRI sequences: Participants will be trained in all functional MRI scan tasks prior to entering the scanner.

Participants will use a mirror and back-projection screen system with a response glove. Participants will be able to view a screen at the back of the scanner via a mirror fixed to the head coil. They will be given a response glove to wear in each hand with buttons located under each finger. Depending on the functional MRI task, participants will be instructed to press one of 4 buttons underneath the middle and index fingers of their left and right hands.

Two functional MRI scans will involve identifying visual targets by pressing the correct button on a response box. For example, in one task, participants will see a series of color words (red, blue, yellow, green) and will be instructed to identify the color in which the word is presented as quickly as possible using response buttons on a key pad. In a second task, the participants will see numbers and will be asked to identify which one is different from the others. In the final task, subjects will view pictures of people and scenes. When viewing the pictures, they will be instructed to either allow themselves to experience any resulting emotions they feel about the picture naturally or to change how they feel by thinking differently about the picture. For example, when viewing a picture of a person in a hospital bed, then they can think of a quick and speedy recovery for that person. This is also called positive re-framing. As some subjects may find the nature of some of the pictures upsetting (e.g., they may feel disgusted if they see a picture of a dirty floor), we will allow them to view sample pictures before the MRI to prepare them. Only subjects who agree to view the pictures after practicing the task will be administered this paradigm during the scan. Participants completing the picture paradigm will also complete the Post-Task Questionnaire which assesses the strategies that were used to change their feelings about the pictures during the task.

An additional functional scan will be taken while the participant is at rest. The participant will be instructed to relax and lie still while looking at a fixation cross on a screen while remaining awake. This scan provides information on the brain networks involved while an individual is at rest.

Participants will be reminded periodically that they can discontinue the MRI protocol at any time. This will be done by a two-way communication system that allows for continuous monitoring and verbal exchanges with the participant throughout the MRI protocol. However, discontinuation of the MRI session may make them ineligible for continuation in the study.

Participants will be asked to provide subjective ratings of valence (1=very unhappy; 9=very happy), arousal (1=very calm; 9=very excited), and control (1=not at all; 9=very) during the MRI protocol. Participants will be familiarized with these rating scales beforehand.

All participants will be given a squeeze ball to press in the event that they want to end the scan for any reason. An alarm will sound in the control room and the technician will stop the scan and remove the participant from the scanner immediately.

###### 1.5.4 POST RANDOMIZATION TASKS:

Actigraph Link will be worn for one week every 6 weeks for each study participant. Measures of exercise self-efficacy are collected during the 3rd week of the intervention protocol.

###### 1.5.5 TIME POINT 2 (6 months):

At 6-months into the exercise sessions, the participants will complete the assessments that were conducted at baseline. These assessments should be completed within a 1 month window; therefore 2 weeks before their midpoint date or 2 weeks after their midpoint date. The EMA measurement will be completed at month 5 to ensure all other key physiological assessments are completed on time.

###### 1.5.6 TIME POINT 3 (12 months – post intervention):

At the 12-month session, participants will complete all the assessments that were done at baseline and 6 months. These assessments should be completed within a 1-month window; therefore 2 weeks before their end date or 2 weeks after their end date. It is encouraged to complete session 1 and session 2 within 2 weeks of the intervention completion. If someone is going on vacation after the intervention, then they may be brought in during their last week of intervention for the 12-month assessments. The EMA assessment will begin at Month 11 to ensure all other key physiological assessments are completed on time.

##### 1.6 RANDOMIZATION:

After the completion of the baseline measures and sessions described above the participants will be randomly assigned to one of two groups. These include (a) an exercise condition that will consist of 150 minutes per week of moderate-to-vigorous intensity exercise, or (b) a PA and health information group. We will use a computer randomization protocol that will incorporate a verification that all session visits have been completed and data entered into the database. We will use a minimization algorithm with equal allocation to one of the two groups. The use of the minimization strategy will ensure treatment balance on the two factors of age at study entry (=<42, >42) and gender. Once randomized, an assigned staff member will contact the participant via phone with the group information. On this contact, the staff member will give information such as date, day, time, and location of the subject's first intervention session.

##### 1.7 TARGETED ENROLLMENT TABLE

| | | | | | | | | | | | | | |
|---|---|---|---|---|---|---|---|---|---|---|---|---|---|
| **Year 1** | | | | | | | | | | | | | |
| | **September** | **October** | **November** | **December** | **January** | **Febuary** | **March** | **April** | **May** | **June** | | | |
| Target (random) | | | planning/procedures development | | | | | 2 | 2 | 2 | | | 6 |
| Enrolled | | | | | | | | | | | | | |
| Randomized | | | | | | | | | | | | | |
| **Year 2** | | | | | | | | | | | | | |
| | **July** | **August** | **September** | **October** | **November** | **December** | **January** | **February** | **March** | **April** | **May** | **June** | |
| Target (random) | 2 | 3 | 3 | 3 | 3 | 3 | 4 | 6 | 7 | 7 | 7 | 5 | 53 |
| Enrolled | | | | | | | | | | | | | |
| Randomized | | | | | | | | | | | | | |
| **Year 3** | | | | | | | | | | | | | |
| | **July** | **August** | **September** | **October** | **November** | **December** | **January** | **February** | **March** | **April** | **May** | **June** | |
| Target (random) | 4 | 4 | 7 | 7 | 5 | 3 | 4 | 7 | 8 | 8 | 8 | 6 | 71 |
| Enrolled | | | | | | | | | | | | | |
| Randomized | | | | | | | | | | | | | |
| **Year 4** | | | | | | | | | | | | | |
| | **July** | **August** | **September** | **October** | **November** | **December** | **January** | **February** | **March** | **April** | **May** | **June** | |
| Target (random) | 6 | 7 | 7 | | | | | | | | | | 20 |
| Enrolled | | | | | | | | | | | | | |
| Randomized | | | | | | | | | | | | | |
| **Year 5** | | | | | | | | | | | | | |
| | **July** | **August** | **September** | **October** | **November** | **December** | **January** | **February** | **March** | **April** | **May** | **June** | |
| | | | | | | | | | | | | | 150 |

#### 1.8 <u>INTERVENTION:</u>

**1.8.1 <u>AEROBIC EXERCISE GROUP:</u>** The purpose of conducting the physical activity intervention is to engage otherwise low active individuals in the recommended weekly amount of moderate-to-vigorous aerobic activity (i.e., 150 minutes). This amount was chosen because it has been shown to promote substantial health benefits among adults, including better heart and brain health.

Participants in the aerobic exercise condition will be scheduled for individual sessions with the goal of obtaining 60 minutes of supervised exercise twice per week and one unsupervised (home-based) exercise session lasting 30 minutes. The aerobic exercise group will engage only in aerobic-based exercise (i.e., no resistance or strength-based exercises) utilizing the facilities and resources available at the location. Walking, jogging, or running on a treadmill will be the encouraged mode of exercise. However, study participants will be allowed to use other types of aerobic exercise equipment. Participants will begin at a lighter intensity (50-60% Heart Rate Reserve or HRR) for 15-20 minutes during the first week of the program and gradually increase their intensity and duration by 5 minutes each subsequent week until they reach 60 minutes of exercise depending on the group (see more below). If the participant elects to increase the intensity at a faster rate and the exercise physiologist considers this safe, then the participant will be allowed to progress at a faster rate. All exercise sessions start and end with 5-10 minutes of stretching for the purpose of warming up and cooling down. Levels of exercise intensity will be prescribed based upon maximal responses during the initial graded exercise treadmill test and then reassessed after the midpoint graded exercise treadmill test. As the participants will be low active at baseline, the prescribed intensity will be 50–60% of the maximum heart rate reserve for weeks one to six and 75–85% for the remainder of the program as long as it is deemed safe by the monitoring exercise physiologist and trainer. In addition to heart rate readings, we will monitor self-reported intensity based on Rating of Perceived Exertion (RPE 6 – 20 scale) with a light intensity considered as the range of 11 – 12 for weeks one to six and then increase to 13 – 14 for remainder of the study. Participants are allowed to go into the vigorous range of 75-85% of HRR as long as it is deemed safe by the lead exercise physiologist and monitoring trainer. Participants that feel comfortable increasing their levels at a more rapid pace will be allowed to do so if the accompanying trainer also agrees that it is safe. Participants in the aerobic group will wear a heart rate monitor and be encouraged to exercise within their target heart rate zone. Heart rate intensity will be measured by a Polar H10 heart rate strap that is worn below their chest. Certified exercise instructors will closely monitor attendance, intensity, frequency, and safety. On-site exercise sessions will occur two days/week for the duration of the intervention and at home for one other day of the week. Compliance to home-based exercise will be monitored by exercise diaries in which participants will record mode, duration, and intensity of exercise (RPE).

Several strategies will be utilized by the exercise trainers to promote adherence and compliance to the exercise intervention, these include the following:

• Consistent and positive feedback in relation to exercise intensity, progress, attendance and duration.
• Foster realistic expectations on behalf of the participant. If expectations are too high, this may lead to a lack of adherence or possible injury by attempting to do too much.
• Flexibility in regard to session times and days. Over a 12-month period, some participants may have major life events (or minor ones) that affect their schedules and availability. It is imperative that we work within reason to accommodate scheduling constraints of participants – the goal is to achieve 150 minutes per week of exercise, and we should accommodate participants if they are willing to engage in this level of activity.
• Individual instruction via face-to-face interaction to raise awareness if the participant is not reaching treatment goals. Review benefits of being more active and work with participants on a plan to reach treatment group goals.
• Use a variety of exercise equipment to reduce boredom and to keep them engaged over the full 12-month period.
• Intervention staff to maintain weekly session logs that track duration, intensity, and type of activity for each session. Exercise trainers can use this information to point out the progress that participant has made while in the program. Example: In week one participant is only able to walk 2.0 mph to reach target heart rate goal for 15 minutes. However, at week 4, they are walking 2.5 mph with 1.0% grade for 30 minutes while maintaining same heart rate goal.
• Continual monitoring of possible barriers to adherence and compliance such as observed physical discomfort or verbal mention of such issues by the participant. It is important that intervention staff create a bond with participants and have open dialogue at each supervised session.
• Follow-up (same day) with study participants who miss session (no show) with no warning to prevent prolonged lapse in attendance.
• Develop strategies with intervention team on a strategy for return to exercise after illness or injury.

**1.8.2 PHYSICAL ACTIVITY & HEALTH INFORMATION GROUP:** The control group will be asked to wear an ActiGraph daily monitor for 7 days every 6 weeks. They will not be given any information on exercise and they will not be asked to change their behavior or exercise patterns.

**1.8.3 EXERCISE COMPLIANCE:** Participants in the study will be completing part of the exercise under supervised conditions and part of the exercise under unsupervised conditions (e.g., home exercise). Monitoring compliance during the supervised exercise is relatively straightforward because we will have trainers monitoring attendance, duration of exercise, perceived exertion, mode of activity, and heart rate intensity throughout the period. However, compliance during the unsupervised sessions (~30 minutes) is inherently more challenging. In this study, we have proposed a two-faceted approach for monitoring compliance during the unsupervised periods. First, within the first few weeks of the intervention, our staff will train participants on the Ratings of Perceived Exertion (RPE) Scale and participants will be instructed to keep weekly diaries of the times, durations, and RPE intensity during the unsupervised exercise that they engage in. These diaries will then be collected on a weekly basis and entered into our database (REDcap). These diaries are a natural component of training since training involves the identification of barriers and approaches to overcoming non-compliance while keeping individuals engaged and continuing to return for the sessions.

In addition to the self-reported diaries of unsupervised exercise, we will have participants wear an Actigraph accelerometer for 1 week every 6 weeks of the study for 12 months (~9 weeks of accelerometer data). We will be able to use these data as objective confirmation that when participants are recording their exercise they are doing the activity at the prescribed intensity and for the prescribed duration. These assessments are important for providing an objective assessment of compliance during the unsupervised exercise periods.

Based on the above, we have operationally defined compliance to the intervention as those participants meeting the prescription of an average of 150 minutes per week of moderate-to-vigorous intensity exercise. Compliance to these prescriptions will be based on the information gathered from the weekly diaries, the supervised exercise sessions, and the accelerometer recordings. We will record on a weekly basis for each participant whether they were compliant or not.

Our analysis plan has proposed an intent-to-treat analysis where all participants, regardless of whether they complete all sessions, will be invited to return for follow-up assessments. However, there will likely be some participants that refuse to return for follow-up assessments or miss a mid-point assessment. “Lost to follow up” will be defined here as missing data that is contributing to the primary outcomes. The reason will be recorded (i.e., missed visit due to traveling, illness). These criteria and documentation will also be applied for any discontinued subjects with the reasons for discontinuation being recorded (i.e., moving out of state).

Regarding the analytical approach, we propose to use all available data in the planned analysis for testing our primary outcome. Since we are examining changes over time we will be able to conduct the analysis as long as there is at least one follow-up assessment (either at the 6-month time point or the 12-month time point). We will assess the missingness of data consistent with the assumptions of the modeling approach described above. That is, we will examine the randomness of missing data and determine if the data is missing completely at random (MCAR) or missing at random (MAR) and use maximum likelihood estimation for computation of unbiased parameters estimates and standard errors. We will not conduct multiple imputation to correct for missing data.

#### 1.9 PARTICIPANT SAFETY:

The safety of study participants will be monitored throughout the study. Study participants will undergo an extensive screening prior to entering the trial to determine that it is safe for them to participate in the intervention. Safety is continuously monitored during the intervention via two supervised sessions per week. We are aware of the potential for serious adverse events to occur with any type of exercise training. As such, the trial will take measures to reduce the risk of an adverse event. All individuals involved in human subject's research, regardless of whether exercise training is involved, are trained and certified in CPR and First Aid. We will have a medical monitor (cardiologist) who will review any exercise ECGs that are flagged for any unusual reading. Included in the exercise stress testing room is an automated external defibrillator (AED), should it be deemed necessary. Lastly, protocols are currently in place to respond to any adverse events that include contacting emergency response personnel and facilitating their arrival to the correct rooms.

The Data Safety Monitoring Board (DSMB), is responsible for monitoring study data for evidence of adverse effects attributed to the intervention. The DSMB also has the authority to recommend changes, stopping the trial at any time, or pausing the trial until problems are resolved. All AEs experienced by the participants during the study (Consent signing until end of study outcome collection) are to be reported during the regularly held DSMB meeting. The eBACH study will track the occurrence of:

- Serious Adverse Events (SAE)
- Unexpected Events
- Unfavorable medical events that occur during intervention sessions

The trial has created two forms for adverse event reporting. These documents have been created based on the belief that trainers and staff should not be responsible for determining whether the event is related to the intervention or whether/how it should be reported to the IRB and DSMB. Only the PI will determine if the event is (a) expected or unexpected, and (b) related to the intervention. Given the frequency of the interactions between subjects and trainers it will be important for trainers and staff to ask if the participant has had any illnesses, injuries, or medication changes since their last supervised exercise session. Based on responses to these questions, regardless of seriousness or relatedness, the trainer will complete a form documenting an adverse event. We have created a brief form for trainers (eBACH Adverse Event Form for Trainers) for this purpose. Trainers or staff would first document the event on this form. This form allows trainers to circle the reported event (or provide an event not listed on the form). This completed document would be sent to the project coordinator who would then speak to the trainer and/or participant to find out more information about the event. The project coordinator would then complete the eBACH Event Evaluation Form based on this communication. This form has several sections with boxes available for documenting the reported severity (mild, moderate, severe), the chronicity (single occurrence, intermittent, persistent), whether the event was resolved or not, and whether it was related to the intervention and expected or unexpected. In particular, these forms are intended to clarify for the investigators, the IRB, and the DSMB, whether the adverse event was serious, whether the adverse event was related to the intervention, and whether it was expected or unexpected. An example of an expected adverse event from the exercise intervention would be muscle soreness which would be listed as a known risk in the IRB. An unexpected adverse event related to the intervention would be if someone falls off a treadmill and injures an ankle. The PI will oversee the determination of these events and if there is any question about whether the event was related to the intervention, its seriousness or expectedness, the event will be discussed at the weekly project meeting. For DSMB purposes, all adverse events that are reported will be recorded and separated based on whether the event was related to the intervention, unexpected, its seriousness, severity, chronicity, and resolution.

#### 1.10 DATA MANAGEMENT:

We appreciate the challenges associated with the organization, execution, management, and analysis of a study of this kind and recognize the importance of frequent communication between staff, students, analysts, and investigators. Protection of subject privacy and safety, meticulous data quality checks, and prudent organization is the bedrock for success of a study like this, which will require a sizeable team/staff.

IT staff will control permissions to the database to ensure proper access and data management by staff, students, and investigators. We will store and link all behavioral and assay data on a HIPPA secure cloud-based server (REDcap).

The data forms in REDcap are similar to the actual paper version of the forms that will be completed by participants and staff. All participant data will be stored and archived on a secure server at the University of Pittsburgh. The server is set for safe mode that allows two copies of each data to be saved on the server. By having the data base on REDCap, all data is secure is case of a natural disaster in which the server would be completely undamaged.

Missed Visits: All outcome collection must be reported and stored in the database. Therefore, if an outcome visit is missed or not collected, a missed visit form must be completed in REDCap. Once a missed visit form is completed, the data management team will indicate in the database that the outcome data for the specific measure is missed to follow up. All missing data in the database will require additional documentation regarding whether the data are "lost to follow up" or have been collected but there was a problem with it.

The missed visit form has three sections that must be completed:

- Time-point of the missed visit
- Outcome that was missed
- Reason the outcome was missed

If a study participant misses multiple outcome measures during the same time point, one missed visit form can be completed if the reason missed is the same for each measure. Thus, staff do not have to complete a new form for each missed outcome for the same time point. If this study participant would also miss post intervention visits, a new form must be generated since this is a different time point.

#### 1.11 QUALITY CONTROL:

There will be multiple layers to the quality control process. First, we will have double entry for all paper-pencil assessment measures that need entered into REDCap to ensure that data is entered correctly. A separate person will review the data for any discrepancies. Before randomization will occur, the study coordinator will check all data for completeness and quality. At which point, the study coordinator will complete the data management quality checklist and give approval for randomization to occur. At weekly meetings the data will be reviewed to identify any issues with QC.

##### 1.11.1 MRI QUALITY CONTROL:

Research associates (RAs) trained in MRI data acquisition and administration of tasks as well as placement of physiological equipment (e.g., blood pressure cuff) will be present at every scan. These RAs will be accompanied by another research staff member to assist in participant instruction and monitoring of physiological data during the scan. A trained MRI technician from the BRIDGE Center will be present to operate the MRI scanner and Siemens software, orienting all field of view maps and ensuring proper slice placement. Data acquired in the MRI scanner will be distributed to the Erickson and Gianaros labs in parallel for simultaneous quality checks and maintenance of copies of original data. Data will enter a quality check pipeline that includes 4 quality control steps: (a) checking of correct image acquisition parameters, (b) an assessment of image quality parameters (i.e., signal-to-noise, contrast-to-noise, etc.), (c) an evaluation of appropriate slice orientation and brain coverage for each sequence, and (d) preprocessing to ensure minimal movement in fMRI scans.

##### 1.11.2 ACTIGRAPHY, FITNESS, EMA, CARDIOVASCULAR QUALITY

**CONTROL**:

All actigraphy, fitness, EMA, cardiovascular assessments, and questionnaires will be interrogated at baseline before randomization to confirm data quality (including means, standard deviations, out of range values, etc.). Quality control assessments will continue on a bi-weekly basis and values will be reported to the Principal Investigator for possible discussion during weekly project meetings.

#### 1.12 RESOURCE SHARING:

##### 1.12.1 DISSEMINATION PLAN:

First, the trial will be registered at ClinicalTrials.gov after formal approval of the protocol by the Data Safety and Monitoring Board, which is expected to be confirmed within the first 6 months of funding. Once the protocol is approved by ClinicalTrials.gov, updates to recruitment and status of the project will be provided to NHLBI every 3 months, or more frequently if requested, until the closure of the study. Finally, results from the primary and secondary aims will be uploaded to ClinicalTrials.gov within 12 months of the completion of the clinical trial. Dr. Erickson has significant experience working with ClinicalTrials.gov in other clinical trials and will be responsible for seeing that these results are updated in a timely fashion.

In addition, participants will be informed of the posting of information to ClinicalTrials.gov as part of the Informed Consent process and will be informed that there will not be any individual data uploaded or any private or confidential information shared on the ClinicalTrials.gov website.

Finally, the University of Pittsburgh has internal policies in collaboration with the Human Research Protection Office (HRPO) to ensure that the registration and result reporting are in compliance with federal policy. Dr. Erickson will work with the Institutional Review Board, the Data Safety and Monitoring Board, and the University HRPO to ensure that these policies are followed.

We will develop an extensive dissemination plan and will publicize the results of the study to practicing clinicians, policy makers, research study participants and the general public by utilizing diverse strategies outlined below. Dissemination will occur through the following techniques: (a) media coverage through press releases and interviews targeted to local and national newspapers, television and radio outlets; (b) production of a research summary document and “facts sheet” targeted to the general public which clearly and concisely summaries the key conclusions of the trial; (c) production of flyers, posters, brochures, and research briefs targeted to broad audiences; (d) study newsletters targeted to study participants; (e) utilizing our study website to share news and research findings to study participants and the public; (f) publishing the results from the trial in high impact and high quality scientific journals within 12 months of closure of the study.

The trial has created a publications policy structure that also provides guidelines to students and investigators about authorship, data analysis and accessibility approaches, etc.

**1.12.2 DATA SHARING:**

We are familiar with the NIH policies regarding data sharing and will comply with local, state, and federal laws, such as the Privacy Rule, a Federal regulation under the Health Insurance Portability and Accountability Act (HIPAA). We will follow the NIH data sharing guide and provide a data sharing plan to be reviewed and approved by the relevant NIA PO. Data sharing will be accomplished using mixed modes, each providing a different level of access. This will include data enclave (controlled, secure environment in which eligible researchers can perform analyses using data resources), researcher's efforts (investigator responds to data requests), and publishing articles in scientific publications. Each of these methods will require investigators to complete data request forms to document aims, procedures, and to ensure that all investigators understand the policies put into place by the study team. We propose to use **REDcap** (Research Electronic Data Capture) as a data enclave for investigators that may request access to certain data. REDcap is a secure web application for building and managing surveys and databases. It allows a streamlined process for rapidly creating and organizing databases and allows for automated exporting of data to Excel and common statistical packages (SPSS, SAS, Stata, R) as well as a built-in project calendar, a scheduling module, ad hoc reporting tools, and other features.

**1.12.3 DATA SHARING AGREEMENT:**

The data-sharing agreement provides for: (1) a commitment to using the data only for research purposes and not to identify any individual human participant; (2) a commitment to securing the data using appropriate computer technology; (3) a commitment to destroying or returning the data after analyses are completed, (4) a commitment to focus on particular aims documented in the agreement, (5) a commitment to work with the Principal Investigator of the project to ensure accuracy of analyses and interpretations, and avoid errors. We will make the data available to outside investigators using a data sharing agreement after publication of the primary aims and at completion of quality control assessments and data organization. Protecting the rights and privacy of human subjects will be our first priority. The final datasets will be de-identified prior to release for sharing.

14 Organization, W. H.

15 NHLBI. <https://www.nhlbi.nih.gov/health/health-topics/topics/phys/benefits>

16 Crichton, G. E., Elias, M. F., Davey, A. & Alkerwi, A. Cardiovascular health and cognitive function: the Maine-Syracuse Longitudinal Study. *PloS one* **9**, e89317, doi:10.1371/journal.pone.0089317 (2014).
17 Kuller, L. H. *et al.* Subclinical Cardiovascular Disease and Death, Dementia, and Coronary Heart Disease in Patients 80+ Years. *Journal of the American College of Cardiology* **67**, 1013-1022, doi:10.1016/j.jacc.2015.12.034 (2016).
18 Nagai, M., Hoshide, S., Ishikawa, J., Shimada, K. & Kario, K. Ambulatory blood pressure as an independent determinant of brain atrophy and cognitive function in elderly hypertension. *Journal of hypertension* **26**, 1636-1641, doi:10.1097/HJH.0b013e3283018333 (2008).
19 Okonkwo, O. C. *et al.* Physical activity attenuates age-related biomarker alterations in preclinical AD. *Neurology* **83**, 1753-1760, doi:10.1212/WNL.0000000000000964 (2014).
20 Raz, N. & Rodrigue, K. M. Differential aging of the brain: patterns, cognitive correlates and modifiers. *Neuroscience and biobehavioral reviews* **30**, 730-748, doi:10.1016/j.neubiorev.2006.07.001 (2006).
21 Rovio, S. *et al.* Leisure-time physical activity at midlife and the risk of dementia and Alzheimer's disease. *Lancet neurology* **4**, 705-711, doi:10.1016/S1474-4422(05)70198-8 (2005).
22 Hallgren, M. *et al.* Exercise, Physical Activity, and Sedentary Behavior in the Treatment of Depression: Broadening the Scientific Perspectives and Clinical Opportunities. *Frontiers in psychiatry* **7**, 36, doi:10.3389/fpsyt.2016.00036 (2016).
23 Pedersen, B. K. & Saltin, B. Exercise as medicine - evidence for prescribing exercise as therapy in 26 different chronic diseases. *Scandinavian journal of medicine & science in sports* **25 Suppl 3**, 1-72, doi:10.1111/sms.12581 (2015).
24 Pemberton, R. & Fuller Tyszkiewicz, M. D. Factors contributing to depressive mood states in everyday life: A systematic review. *Journal of affective disorders* **200**, 103-110, doi:10.1016/j.jad.2016.04.023 (2016).
25 Murtagh, E. M. *et al.* The effect of walking on risk factors for cardiovascular disease: an updated systematic review and meta-analysis of randomised control trials. *Preventive medicine* **72**, 34-43, doi:10.1016/j.ypmed.2014.12.041 (2015).
26 Colcombe, S. J. *et al.* Aerobic exercise training increases brain volume in aging humans. *The journals of gerontology. Series A, Biological sciences and medical sciences* **61**, 1166-1170 (2006).
27 Voss, M. W. *et al.* Plasticity of brain networks in a randomized intervention trial of exercise training in older adults. *Frontiers in aging neuroscience* **2**, doi:10.3389/fnagi.2010.00032 (2010).
28 Voss, M. W. *et al.* The influence of aerobic fitness on cerebral white matter integrity and cognitive function in older adults: results of a one-year exercise intervention. *Human brain mapping* **34**, 2972-2985, doi:10.1002/hbm.22119 (2013).
29 Chaddock-Heyman, L. *et al.* The effects of physical activity on functional MRI activation associated with cognitive control in children: a randomized controlled intervention. *Frontiers in human neuroscience* **7**, 72, doi:10.3389/fnhum.2013.00072 (2013).

30 Chaddock-Heyman, L. *et al.* Aerobic fitness is associated with greater hippocampal cerebral blood flow in children. *Dev Cogn Neurosci* **20**, 52-58, doi:10.1016/j.dcn.2016.07.001 (2016).
31 Chaddock, L. *et al.* A neuroimaging investigation of the association between aerobic fitness, hippocampal volume, and memory performance in preadolescent children. *Brain Res* **1358**, 172-183, doi:10.1016/j.brainres.2010.08.049 (2010).
32 Thomas, A. G. *et al.* Multi-modal characterization of rapid anterior hippocampal volume increase associated with aerobic exercise. *NeuroImage* **131**, 162-170, doi:10.1016/j.neuroimage.2015.10.090 (2016).
33 Nunley, K. A. *et al.* Physical activity and hippocampal volume in middle-aged patients with type 1 diabetes. *Neurology* **88**, 1564-1570, doi:10.1212/WNL.0000000000003805 (2017).
34 Sandroff, B. M., Johnson, C. L. & Motl, R. W. Exercise training effects on memory and hippocampal viscoelasticity in multiple sclerosis: a novel application of magnetic resonance elastography. *Neuroradiology* **59**, 61-67, doi:10.1007/s00234-016-1767-x (2017).
35 Chaddock-Heyman, L. *et al.* Higher cardiorespiratory fitness levels are associated with greater hippocampal volume in breast cancer survivors. *Frontiers in human neuroscience* **9**, 465, doi:10.3389/fnhum.2015.00465 (2015).
36 Raji, C. A. *et al.* Longitudinal Relationships between Caloric Expenditure and Gray Matter in the Cardiovascular Health Study. *Journal of Alzheimer's disease : JAD* **52**, 719-729, doi:10.3233/JAD-160057 (2016).
37 Voss, M. W. *et al.* Neurobiological markers of exercise-related brain plasticity in older adults. *Brain, behavior, and immunity* **28**, 90-99, doi:10.1016/j.bbi.2012.10.021 (2013).
38 Burdette, J. H. *et al.* Using network science to evaluate exercise-associated brain changes in older adults. *Frontiers in aging neuroscience* **2**, 23, doi:10.3389/fnagi.2010.00023 (2010).
39 Colcombe, S. J. *et al.* Cardiovascular fitness, cortical plasticity, and aging. *Proceedings of the National Academy of Sciences of the United States of America* **101**, 3316-3321, doi:10.1073/pnas.0400266101 (2004).
40 Erickson, K. I. *et al.* Exercise training increases size of hippocampus and improves memory. *Proceedings of the National Academy of Sciences of the United States of America* **108**, 3017-3022, doi:10.1073/pnas.1015950108 (2011).
41 Rovio, S. *et al.* The effect of midlife physical activity on structural brain changes in the elderly. *Neurobiol Aging* **31**, 1927-1936, doi:S0197-4580(08)00366-7 [pii] 10.1016/j.neurobiolaging.2008.10.007 (2010).
42 Dunn, A. L. *et al.* Comparison of lifestyle and structured interventions to increase physical activity and cardiorespiratory fitness: a randomized trial. *Jama* **281**, 327-334 (1999).
43 Fritz, M. S. & Mackinnon, D. P. Required sample size to detect the mediated effect. *Psychological science* **18**, 233-239, doi:10.1111/j.1467-9280.2007.01882.x (2007).
44 Gibbons, R. D. & Hedeker, D. Random effects probit and logistic regression models for three-level data. *Biometrics* **53**, 1527-1537 (1997).

### Amendments in Study Protocol

Given the continued challenges of navigating the COVID-19 pandemic, we requested to modify our recruitment/randomization accrual and targeted sample size for the study titled “Exercise on Brain and Cardiovascular Health (eBACH)”, Project 3 of P01-HL040962, on September 28th, 2021. These recommended changes were approved by the meeting of the Data Safety and Monitoring Board on September 2nd, 2021. The details of this requested change are described below.

- Our original targeted sample size of 150 was re-evaluated by our study statistician (Dr. Chaeryon Kang, Department of Biostatistics, University of Pittsburgh) with a focus on the minimal sample size needed to be sufficiently powered to test our primary hypotheses. Based on prior literature described in the grant application, we estimated effect sizes between .5 and .6 (Cohen’s d) for changes in brain volume resulting from an exercise intervention. Using mixed effects modeling of the data in an intent-to-treat approach, we will have between 68-83% power to test our primary aim (hippocampal volume change) with 120 participants, which accounts for a possible 20% attrition rate. Similar effect sizes for the effects of exercise on various cardiovascular disease biomarkers have been reported (between .58 and .6). Therefore, a sample of 120 would provide us >80% power even with 20% attrition to test our hypotheses about the effects of exercise on cardiovascular disease biomarkers and mediators. In addition, we will be sufficiently powered (>80%) to test several of our secondary aims (e.g., depressive symptoms and functional brain connectivity) with a targeted sample of 120. In sum, we have determined that we will have sufficient power to test our primary aims with a reduction in the targeted sample size to 120. What is more at risk with a reduction in sample size is the statistical power for mediation analyses to test our tertiary aims (i.e., to use structural equation modeling approaches for examining mediators between changes in brain and changes in cardiovascular health markers). As such, we propose to move forward by conducting these tertiary aims on an exploratory basis.
- The power estimations described above include an attrition rate of 20%, but our current attrition rate is <10% for all participants that were not disrupted due to COVID. This suggests that the power estimations described above are likely conservative estimates.
- Our current exercise adherence and compliance rates are >90% indicating that we are successful at getting and keeping participants physically active. This is important because low adherence and compliance have a direct effect on our ability to test our hypotheses (i.e., if participants are not
- adherent then we would be unable to test our aims). In short, the exercise sessions and intervention are going well which supports our ability to test our primary and secondary aims with a smaller than originally proposed sample.
- The modified accrual rates described in our letter in December 2020 were overly optimistic at the time and proposed in view of the forthcoming public availability of vaccines and potential normalization of research conduct prior to the rise of SARS-CoV-2 variants. Unfortunately, the year 2021 in the Pittsburgh region has been marred by unanticipated challenges including variable vaccine hesitancy rates across different districts, the rise of the delta and other variants, continued illnesses

and hospitalizations, need for quarantine, reduced density of staff and participants in buildings, school and day care cancellations, etc. (see more on this below). As such, we propose to modify our accrual rates by taking advantage of a 1 year no cost extension (which was not included in our adjusted targets in December of 2020). We have worked closely with our budget office to confirm that we will have sufficient funds for a no cost extension and reducing our proposed targeted sample from 150 to 120 (30 participants or 60 sessions) will also reduce additional costs and time. This new accrual rate is more feasible given current trends in the COVID pandemic.

- There are two primary bottlenecks that we have experienced related to our lower-than-expected accrual rate. First, our study team has experienced recent turnover in staff that were hired for recruitment and screening. This was noted by the study PI and DSMB members at the September 2nd, 2021 meeting. To address this issue, we have submitted hiring materials to the University of Pittsburgh Human Resources office to hire a new staff member who will be dedicated to advertising, screening, and recruitment for the study. We expect this individual to start in October 2021. In the interim, we will immediately have staff from other Projects and Cores assist in recruitment and screening efforts. Secondly, we have had an unprecedented number of session cancellations and participants withdrawing from the study during baseline assessment sessions (before randomization). As such, our screening-to-randomization ratio is lower than anticipated which requires the study team to screen more individuals to randomize one person to the intervention. This behavior (session cancellations and withdrawing before randomization) has been observed across several ongoing studies and can be attributed to issues related to the pandemic described above (e.g., need for quarantine). We are currently working through methods to accommodate missed sessions or make participation in the trial more flexible and accommodating to the needs of the participants.
- The DSMB members noted on September 2nd, 2021 that the rate of minority recruitment is lower than anticipated. We are taking active steps to improve this rate including community advertisements in addition to our marketing strategy that we have used up until now. Dr. Gianaros has also been attending local farmer's markets to recruit within diverse neighborhoods in Pittsburgh, and we have a forthcoming meeting with an elected Councilperson in the Wilkinsburg district of Pittsburgh to formulate a community-based recruitment effort to reach out to a more diverse sample to meet enrollment targets. Once our new staff are hired and trained, we will be actively implementing these strategies to enhance minority recruitment. If necessary, we will pause the recruitment of non-Hispanic whites into the trial in order to focus on increasing enrollment of underrepresented minority groups.
- Lastly, as noted in the attached and updated accrual plan approved by the DSMB, we can report that we are currently 4 people lower than our randomization target as of this writing. Hence, 57 of 61 people have been randomized, which reflects 93% of the new DSMB approved randomization goal for this point in time.
