## Supplement 2_Supplemental Material for "Fitness and Exercise Effects on Brain Age: A Randomized Clinical Trial"

**Supplementary Content**

**Supplemental Methods.**

**Supplemental Results.**

**References.**

**Supplemental Tables and Figures.**

**Figure S1.** Brain-predicted age and brain-PAD at baseline and 12-month follow-up.
**Figure S2.** Intervention effect with changes in marginal mean values of (A) brain-PAD and (B) $VO_{2peak}$ from baseline to 12 months by ITT allocation, including only completers.
**Figure S3.** Association of changes in $VO_{2peak}$ with changes in brain-PAD in (A) all samples, (B) aerobic exercise group, and (C) control group.
**Table S1.** Regression analyses examining the association between CRF and brain-PAD at baseline.
**Table S2.** Descriptive mean values and SDs of brain-PAD, CRF, and other biological measurements by ITT allocation and study time point.
**Table S3.** Estimated marginal means for brain-PAD, CRF, and biological measurements comparing the intervention and control arms, including only completers.
**Table S4**. Adverse events during the eBACH trial.

### Supplemental Methods

#### Study Design and Participants

In this single-center, parallel-armed RCT (ClinicalTrials.gov: NCT03841669), healthy adults aged 26-58 years were recruited for participation via multiple methods, including (a) mass electronic and print mailings to Allegheny County residents in Pennsylvania, USA; (b) radio, electronic, and print advertisements in public places; and (c) direct solicitation from University of Pittsburgh Research Participant Registries. Individuals who responded to recruitment solicitations were screened by phone, and those who remained interested in volunteering were again screened by an in-person medical history interview prior to further testing. The study protocol and informed consent process were approved by the University of Pittsburgh Institutional Review Board (IRB ID: 19020218). All participants provided written informed consent before enrollment. More details are available in the published protocol.[1]

Participants were eligible if they reported no history or presence of chronic psychotic illness (e.g., schizophrenia, bipolar disorder) or neurological disorder (e.g., Parkinson's disease, dementia, mild cognitive impairment (MCI)), no current treatment for cancer or treatment in the past year, Type 1 diabetes, or current use of prescribed blood pressure medication.  Before enrollment, all participants self-reported exercise for fewer than 100 minutes per week and achieved a $VO_{2max}$ percentile less than 75 based on the ACSM Guidelines for Exercise testing (9th and 10th edition). Additionally, participants were required to be free of any MRI contraindications (e.g., metallic implants unsafe for scanning).

#### Randomization and Blinding

Participants were randomly assigned to either a moderate-to-vigorous intensity aerobic exercise condition or to a health information control condition for a 12-month intervention period. Randomization was conducted using stratified block randomization on a web-based system through the Research Electronic Data Capture (REDCap) randomization module by the study biostatistician (CK) to ensure unbiased allocation of participants to the two intervention groups by age and sex. All investigators and staff involved in data collection were blinded to group assignment. Only staff involved in the implementation of the exercise intervention, scheduling of sessions, and study coordination were unblinded to group assignment. No reports of incidental unblinding occurred.

#### Exercise Intervention and Control

##### Aerobic Exercise Condition

Participants engaged in supervised exercise two times a week for 120 minutes per week (60 minutes per session) in a laboratory setting at the University of Pittsburgh along with home-based exercise to achieve the prescribed 150 minutes of exercise per week. Exercise duration progressed from 75 minutes in week one to the final goal of 150 minutes starting with week 4. Participants were encouraged to walk, jog, or run on a treadmill, as well as to record the use of other types of aerobic exercise equipment such as bikes, elliptical machines, stair climbers, and rowers. Levels of exercise intensity were prescribed based on maximal responses during the initial graded exercise test (GXT). For weeks 1-6, the prescribed intensity was 50%-60% of the maximum heart rate reserve (HRR). For the remainder of the intervention, participants increased their intensity to 60%-75% of HRR. as long as it was deemed safe by the monitoring exercise physiologist and trainer, reaching the prescribed goal of 150 min/week. Participants wore a heart rate monitor (Polar A370 or Unite watch, Polar USA, Lake Success, NY) during all supervised sessions to ensure that exercise was occurring within the target heart rate zone.  Exercise attendance, intensity, frequency, and safety were monitored by certified exercise instructors. Compliance with home-based exercise was monitored by exercise diaries and heart rate monitors. Participants were instructed to record all unsupervised exercise sessions and average heart rate during the exercise as indicated on the heart rate monitors. All exercise sessions consisted of 5-10 minutes of warm-up and cool-down periods.

During pandemic-related shutdowns in 2020, exercise trainers and other staff remained in frequent contact with participants and asked all participants to continue to record all exercise behaviors at home. Heart rate monitors were used to record exercise intensity during home exercise sessions under the guidance of exercise physiologists. As soon as University facilities re-opened, we encouraged all participants to return to supervised exercise as long as it was safe and feasible for them, given family and illness constraints. As a result, 89.1% of participants chose to return to the supervised exercise.

##### Control Group

Participants assigned to the health information control group were asked not to change their behavior or exercise patterns and were encouraged to perform activities as they normally would. They were provided information about the health benefits of engaging in physical activity and were contacted approximately every 6 weeks by the investigative team so as to remain in contact with participants in both groups. Participants in the control group wore an Actigraph monitor for 7 days every 6 weeks, such that any change in sedentary behavior or PA was monitored.

#### Physiological and Biological Measures

##### Cardiorespiratory Fitness Testing

Cardiorespiratory aerobic capacity was measured through a graded exercise test (GXT) (TrueOne 2400, ParvoMedics, Salt Lake City, UT) using a modified Balke Protocol [2] at baseline and at the 12-month post-intervention assessment. We conducted a warm-up (walking on a motor-driven treadmill at a constant speed from 2.0 to 3.5 mph) followed by an incremental protocol (i.e., two-minute stages with a 2% increase in incline at each stage). The test speed was self-selected by the participant in consultation with the exercise physiologist at increments of 0.5 mph within the 2.0–4.0 mph range.

Heart rate was continuously monitored via a 12-lead electrocardiogram (NASIFF CardioSuite ECG cart) along with blood pressure readings (Tango 2, SunTech Medical) and Rating of Perceived Exertion (RPE) every two minutes. $VO_{2Peak}$ was the highest $VO_2$ value recorded during the maximal test using 15-second timed interval averaging. The criteria for achieving peak oxygen uptake ($VO_{2peak}$) were determined based on the American College of Sports Medicine (ACSM) criteria [3]: (i) plateau in $VO_2$ between two or more workloads (increase less than 0.15 L/min or 2.0 ml/kg/min during the last minute of corresponding workloads); (ii) respiratory exchange ratio ≥1.10; (iii) heart rate within 10 beats of age-predicted maximal heart rate (220-age); and (iv) Rating of RPE ≥17. Each test was administered by an exercise physiologist.

##### Stadiometer Measurements

Height and weight were recorded before the GXT at both baseline and post-intervention. Body weight was measured with a calibrated digital scale to the nearest 0.1kg, wearing light clothing, empty pockets, and without shoes. Height was assessed using a calibrated stadiometer (wall-mounted) graduated in centimeters with a horizontal measuring block (or fixed angle), to the nearest 0.1 cm. We calculated body mass index (BMI) by dividing weight (kg) and height ($m^2$).

##### Body Composition

At baseline and at follow-up (post-intervention), all participants' waist circumferences were measured at the level of the umbilicus to the nearest 1/2 centimeter at end expiration. Height was measured by a vertical-mounted tape measure (with shoes off), and weight was measured in kg using a bioelectrical impedance device (Body Composition Analyzer, model TBF-410, Tanita Corp., IL). The Tanita device also provided a measurement of percentage of body fat at the time of measurement.

##### Blood Pressure

Seated resting blood pressures (BP) were obtained with an Omron IntelliSense© BP Monitor (model HEM-907XL, Omron Healthcare Inc). At baseline and post-intervention, three BP readings were taken after five minutes of seated rest. The average of the final 2 of the 3 readings was used to compute resting systolic (SBP) and diastolic blood pressures (DBP) for analysis. Mean arterial pressure (MAP) was further calculated from SBP and DBP.

##### Blood Assays

Plasma levels of BDNF were measured from fasting blood samples collected in citrate collection tubes (BD Vacutainer, NJ) and processed and stored in an ultra-cold freezer for analysis in batches. Both samples collected from each participant were assayed on the same cartridge. All samples were run in triplicate by automated sandwich ELISA on a Simple Plex Human BDNF Cartridge (# SPCKB-PS-000389) using the antibody-based Ella™ system (ProteinSimple, Bio-techne, MM, USA). Assays were run according to the kit protocol.

#### Image Acquisition and Preprocessing

Participants underwent MRI scanning at baseline and 12-month follow-up on a Siemens Prisma 3T scanner with a 64-channel head coil. T1-weighted images were used for brain age estimation and were acquired using the following parameters: Magnetization Prepared Rapid Acquisition of Gradient Echo (MPRAGE) sequence with 1 mm isotropic resolution, 176 slices, 256x256 matrix size, 9° flip angle, repetition time (TR)=2300ms, inversion time (TI)=900ms, and echo time (TE)=1.99ms.

MPRAGE images were submitted to the Computational Anatomy Toolbox (CAT12) implemented in the Statistical Parametric Mapping 12 tool (SPM12) to derive a quantitative image quality rating (IQR) indicating the quality of each T1-weighted image.[4] This image quality metric reflects an aggregate composite of noise, bias, and resolution on a percentage scale, with higher values (100% maximum) indicating better image quality.

#### Brain Age Estimation

Brain age estimation was performed on T1-weighted images using brainageR (v2.1), an open-access software for generating brain-predicted age (github.com/james-cole/brainageR).[5] The pipeline first initiates voxel-level preprocessing using the SPM12 toolbox. This includes segmenting the raw T1-weighted images into gray matter (GM), white matter (WM), and cerebrospinal fluid (CSF), and non-linear spatial normalization to a template image using SPM12's DARTEL (Diffeomorphic Anatomical Registration using Exponentiated Lie Algebra) toolbox. Images were resampled using a voxel size of 1.5 mm and smoothed with a Gaussian spatial smoothing kernel of 4mm at full-width at half-maximum (FWHM). Probabilistic tissue maps were visually inspected to ensure the quality of the segmentation. Normalised tissue maps were loaded to R, converted to vectors, masked using a 0.3 threshold from the mean image template based on the brainageR model training dataset, and then combined.

BrainageR had been pre-trained to predict brain age from normalised brain volumetric maps of 3377 healthy adults (aged 18-92 years) from seven publicly available datasets using a Gaussian Process Regression model.[6] Using principal component analysis, the top 435 principal components capturing 80% of the variance in brain volumes were retained. The pre-trained brainageR model was applied to the vectorized and masked images in the current study to estimate a brain-predicted age score for each of our study participants at each time point.

Brain-predicted age difference (brain-PAD) was further calculated as the deviations of predicted brain age from chronological age. A positive brain-PAD (i.e., older brain age relative to chronological age) suggests an advanced brain age, while a negative brain-PAD (i.e., younger brain age relative to chronological age) represents preserved brain age. To account for a potential age bias on brain age estimates[7,8], we include chronological age as a covariate in the statistical models (refer to *Statistical Analysis*).

#### Statistical Analysis

The mean and standard deviation (SD) were used to describe continuous data, and frequency and percentage were used to summarize categorical data. Baseline demographic characteristics and $VO_{2peak}$ were compared between the aerobic exercise and control groups using the two-samples Wilcoxon rank sum test and Chi-square test for continuous and categorical variables, respectively. Pearson's correlations were conducted to examine associations between brain-predicted age and chronological age and to evaluate for the potential age bias in brain-PAD [7,8].

To investigate the linear association between CRF and brain-PAD at baseline, we used a multiple linear regression model with $VO_{2peak}$ as a predictor and baseline brain-PAD as the dependent variable. We included chronological age, sex, years of education, and BMI as covariates in the model. Previous studies have shown that brain age estimated from the brainageR algorithm is affected by noise and motion artifacts,[9] so the IQR generated from CAT12 was also included as a covariate. Statistical inferences were conducted at the significance level of 0.05 ($p < 0.05$).

We assessed the effect of the exercise intervention on brain-PAD and CRF separately using linear mixed modeling (LMM). The LMM models included the fixed effect for the main effect of treatment (12-month aerobic exercise, control), time (baseline, month 12), and their interaction. We also included a random intercept for individual participants to account for the within-individual correlation among repeated measures at baseline and 12 months. For brain-PAD, we adjusted for randomization factors (baseline chronological age, sex), years of education, BMI, and IQR. For CRF, we controlled for covariates including age, sex, years of education, and BMI. LMM allowed us to model the changes in brain age and CRF as a function of both time and group while also including potential confounding variables. The LMM also considers all available data points, which conforms to the intention-to-treat (ITT) analytical framework. Completers secondary analyses were further performed for testing the intervention effect.

A prerequisite for testing mediation using a statistical mediation framework is that the changes in outcome must be correlated with the changes in the mediator. As such, we first tested the relationship between changes in brain-PAD and changes in CRF, body mass, blood pressure, and BDNF with a series of multiple linear regression models, controlling for chronological age at baseline, sex, and years of education. Considering that the changes in those measurements of interest were not universal and individuals in the control group may also experience meaningful changes, we assessed the associations collapsing across both groups of participants and solely in the aerobic exercise group, respectively. Unstandardized B-values, standardized β-values, and p-values were reported for all regression models.

All analyses were performed using R software (Version 4.2.1).

#### Supplemental Results

##### Missing Data

Of the 130 randomized participants, 129 successfully completed baseline MRI scans, and 81 successfully completed MRI scans at both baseline and 12-month follow-up. The number of missing data points from the original randomized sample were as follows: Exercise Group: N = 23; Control Group: N = 26. A chi-square of the completion ratio between groups was not significant ($p = 0.68$), suggesting the proportion of missing data did not differ by group assignment. Moreover, a comparison of the subsample who did not successfully complete the follow-up MRI to the sample who completed both baseline and follow-up MRI scans revealed no differences in age, percentage of females, or BMI (all $p > 0.05$). However, missingness of the follow-up MRI scans was significantly associated with education levels. Participants who completed 12-month MRI scans had higher education than those with missing MRI scans at 12 months ($t = 2.476$, $p = 0.015$). Based on these associations, our primary regression models were adjusted for the effect of education, assuming missing data was missing at random (MAR), including covariate-dependent missing completely at random (MCAR) as a special case of MAR.

##### Brain-predicted Age and Brain-PAD

When examining the entire sample, the brain-predicted age was 42.22±9.99 years at baseline and 42.54±10.10 years at the 12-month follow-up (Figure S1A). As expected, there was a significant correlation between chronological age and brain-predicted age at both baseline ($r(127)=0.823$, $p<0.001$) and 12 months ($r(79)=0.819$, $p<0.001$; see Figure S1B).

The baseline and follow-up brain-PAD are summarized in Figure S1C. As shown, brain-PAD was 0.97±5.93 years at baseline and 0.55±6.03 years at the 12-month follow-up. We evaluated brain-PAD for age bias as the predicted brain age may be underestimated for older individuals and overestimated for younger individuals [7,8]. Consistent with this, we found a significant correlation between brain-PAD and chronological age (baseline: $r(127)=-0.294$, $p< 0.001$; 12-month: $r(79)=-0.333$, $p=0.002$; Figure S1D).

**Figure S1. Brain-predicted age and brain-PAD at baseline and 12-month follow-up.**

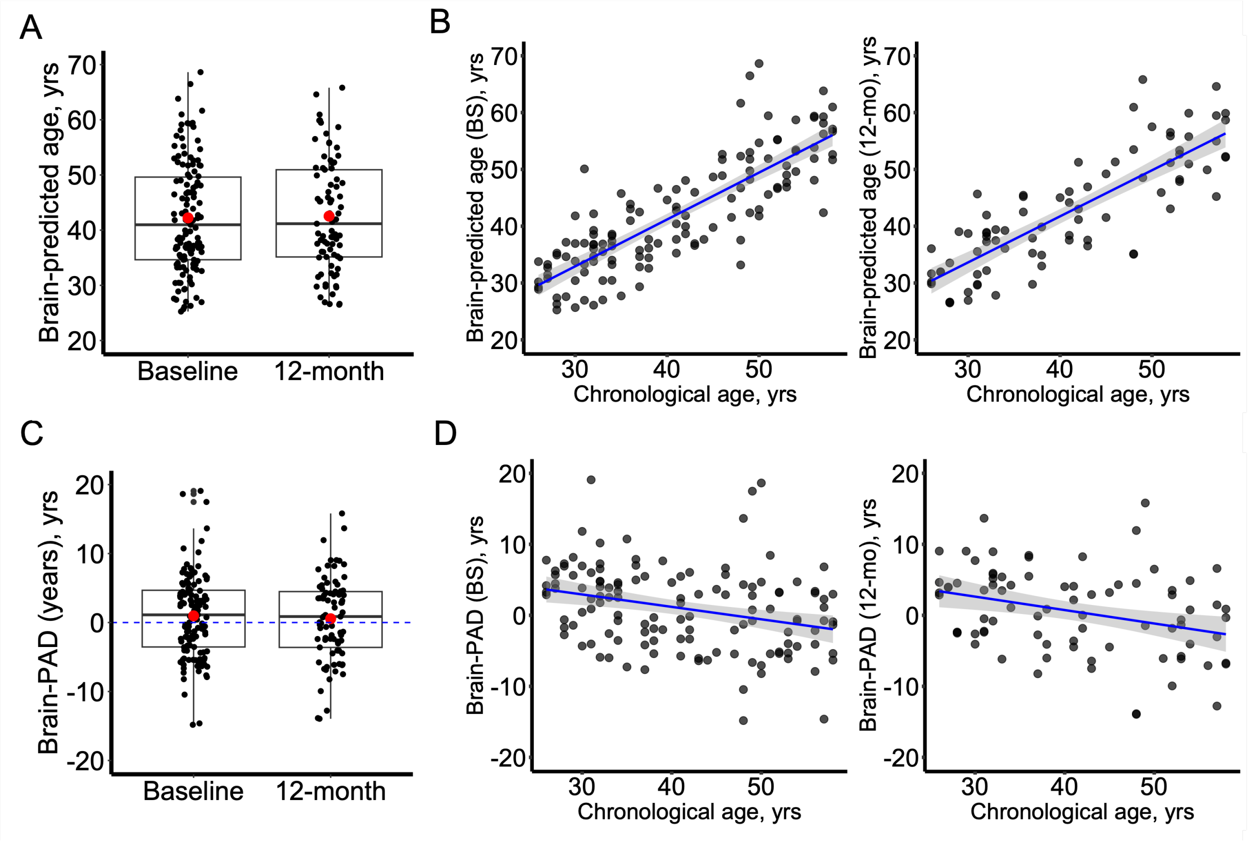


Panel A shows the distribution of brain-predicted age; mean values are indicated by the red dot. Panel B shows the correlation between brain-predicted age and chronological age at baseline (left) and 12-month follow-up (right). Panel C shows the distribution of brain-PAD; mean values are indicated by the red dot. Panel D shows the correlation between brain-PAD and chronological age at baseline (left) and 12-month follow-up (right).
Shaded areas reflect 95% CIs.

**Figure S2. Intervention effect with changes in marginal mean values of (A) brain-PAD and (B) $VO_{2peak}$ from baseline to 12 months by ITT allocation, including only completers.**

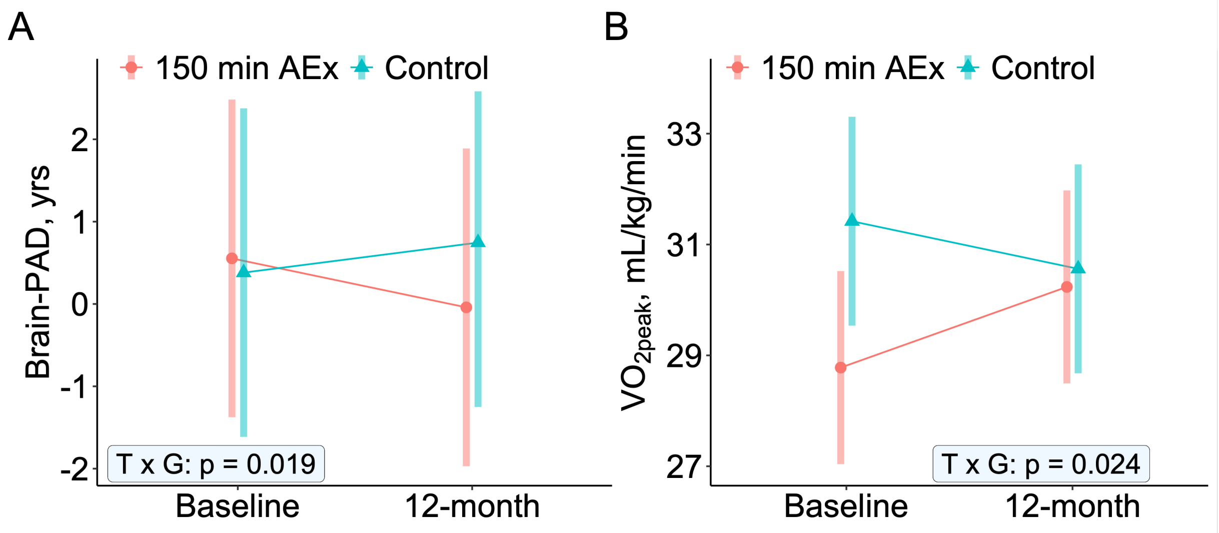


Error bars indicate 95% CIs. T x G stands for the Time-by-Group interaction.

**Figure S3. Association of changes in $VO_{2peak}$ with changes in brain-PAD in (A) all samples, (B) aerobic exercise group, and (C) control group.**

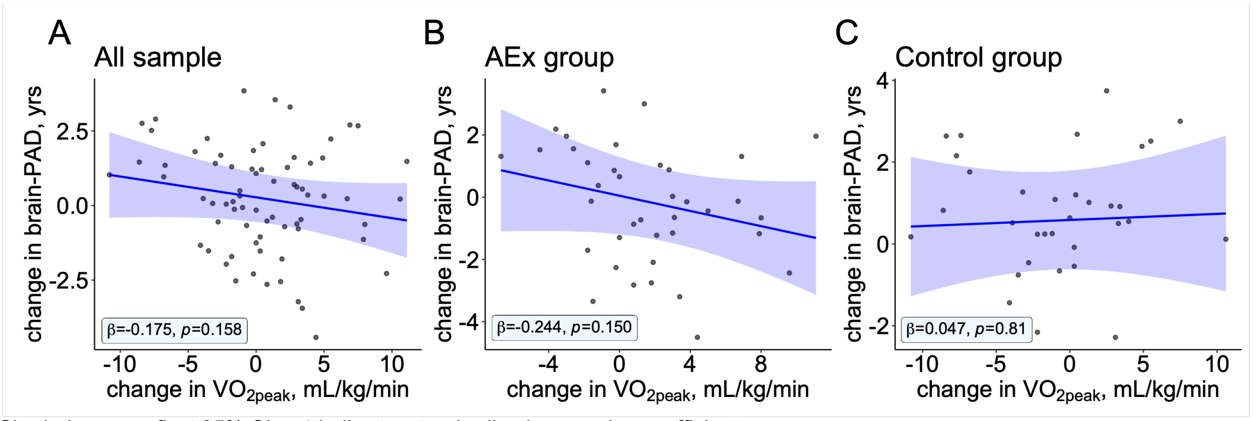


Shaded areas reflect 95% CIs. $\beta$ indicates standardized regression coefficient.

**Table S1. Regression analyses examining the association between CRF and brain-PAD at baseline.**

| Predictors | B | $\beta$ | t | p |
|---|---|---|---|---|
| $VO_{2peak}$, mL/kg/min | -0.263 | -0.309 | -2.549 | 0.012 |
| Age, y | -0.233 | -0.391 | -4.183 | <.001 |
| Sex[a] | 0.160 | 0.013 | 0.133 | 0.895 |
| Education, y | -0.129 | -0.064 | -0.729 | 0.467 |
| BMI, kg/$m^2$ | -0.057 | -0.063 | -0.559 | 0.577 |
| IQR | 4.197 | 0.012 | 0.121 | 0.904 |
| | | $R^2$=0.156, p=0.002 | | |

[a] Female=0; Male=1

B indicates unstandardized regression coefficient, and $\beta$ indicates standardized regression coefficient.

#### Table S2. Descriptive mean values and SDs of brain-PAD, CRF, and other biological measurements by ITT allocation and study time point.

| Variable | Total Sample | | 150min AEx | | Control | |
|---|---|---|---|---|---|---|
| | Baseline mean (SD) | 12-month mean (SD) | Baseline mean (SD) | 12-month mean (SD) | Baseline mean (SD) | 12-month mean (SD) |
| Brain-PAD, y | 0.97 (5.93) | 0.55 (6.03) | 0.73 (5.28) | 0.15 (5.11) | 1.19 (6.53) | 0.96 (6.89) |
| Number, n | 129 | 81 | 63 | 41 | 66 | 40 |
| *CRF* | | | | | | |
| Number, n | 130 | 75 | 64 | 40 | 66 | 35 |
| $VO_{2peak}$, mL/kg/min | 28.57 (6.95) | 29.41 (6.80) | 26.85 (6.60) | 29.26 (6.21) | 30.23 (6.93) | 29.59 (7.50) |
| *Body composition* | | | | | | |
| Number, n | 116 | 80 | 57 | 42 | 59 | 38 |
| BMI, kg/$m^2$ | 28.59 (6.63) | 28.29 (6.14) | 28.97 (6.47) | 28.99 (6.60) | 28.21 (6.81) | 27.52 (5.58) |
| Body fat, % | 33.35 (10.1) | 33.18 (9.94) | 34.17 (9.43) | 33.66 (9.04) | 32.57 (10.73) | 32.65 (10.95) |
| WC, cm | 99.44 (16.39) | 98.19 (15.55) | 100.23 (15.84) | 98.51 (15.68) | 98.69 (17.00) | 97.84 (15.60) |
| *Blood pressure* | | | | | | |
| Number, n | 115 | 79 | 56 | 41 | 59 | 38 |
| SBP, mm Hg | 119.69 (13.98) | 119.46 (13.07) | 122.93 (16.27) | 121.73 (14.64) | 116.61 (10.64) | 117.00 (10.80) |
| DBP, mm Hg | 78.51 (9.52) | 77.19 (10.38) | 79.27 (10.26) | 78.68 (11.67) | 77.80 (8.80) | 75.58 (8.65) |
| MAP, mm Hg | 95.67 (10.69) | 94.66 (10.83) | 97.38 (12.10) | 96.43 (12.49) | 94.06 (8.96) | 92.74 (8.45) |
| *Blood biomarker* | | | | | | |
| Number, n | 112 | 79 | 56 | 41 | 56 | 38 |
| log BDNF, pg/ml | 6.37 (0.79) | 6.60 (0.87) | 6.35 (0.84) | 6.77 (0.85) | 6.40 (0.76) | 6.41 (0.87) |

Abbreviations: Brain-PAD, brain-predicted age difference; CRF, cardiorespiratory fitness; BMI, body mass index; WC, waist circumference; SBP, systolic blood pressure; DBP, diastolic blood pressure; MAP, mean arterial pressure; BDNF, brain-derived neurotrophic factor

#### Table S3. Estimated marginal means for brain-PAD, CRF, and biological measurements comparing the intervention and control arms, including only completers.

| Variable | 150min AEx | Control | Difference between arms [b] | |
|---|---|---|---|---|
| | mean (SE) [95% CI] [a] | mean (SE) [95% CI] [a] | mean (SE) [95% CI] | p |
| Brain-PAD, y | -0.60 (0.28) [-1.15, -0.04] | 0.37 (0.29) [-0.21, 0.94] | -0.96 (0.40) [-1.74, -0.18] | **0.019** |
| *CRF* | | | | |
| $VO_{2peak}$, mL/kg/min | 1.46 (0.69) [0.09, 2.82] | -0.86 (0.73) [-2.32, 0.60] | 2.31 (1.01) [0.34, 4.28] | **0.024** |
| *Body composition* | | | | |
| BMI, kg/$m^2$ | 0.30 (0.20) [-0.09, 0.69] | 0.50 (0.21) [0.09, 0.91] | -0.20 (0.28) [-0.75, 0.36] | 0.488 |
| Body fat, % | 0.18 (0.41) [-0.65, 1.00] | 0.58 (0.43) [-0.28, 1.44] | -0.40 (0.60) [-1.57, 0.77] | 0.501 |
| WC, cm | 0.60 (0.93) [-1.25, 2.45] | 2.51 (0.97) [0.58, 4.44] | -1.9 (1.34) [-0.43, 0.72] | 0.159 |
| *Blood pressure* | | | | |
| SBP, mm Hg | -2.78 (1.79) [-1.56, 0.12] | 1.38 (1.84) [0.75, 0.45] | -4.17 (2.56) [-9.19, 0.85] | 0.108 |
| DBP, mm Hg | -1.22 (1.47) [-4.15, 1.72] | 0.08 (1.15) [-2.94, 3.09] | -1.29 (2.11) [-5.43, 2.85] | 0.543 |
| MAP, mm Hg | -2.00 (1.50) [-4.99, 0.99] | 0.49 (1.54) [-2.59, 3.56] | -2.49 (2.15) [-6.70, 1.72] | 0.251 |
| *Blood biomarker* | | | | |
| log BDNF, pg/ml | 0.21 (0.15) [-0.09, 0.51] | -0.09 (0.16) [-0.40, 0.22] | 0.30 (0.22) [-0.12, 2.72] | 0.167 |

[a] Differences were calculated as values at 12 months minus baseline.
[b] Differences between arms were calculated as 150min AEx group minus control group.
Abbreviations: Brain-PAD, brain-predicted age difference; CRF, cardiorespiratory fitness; BMI, body mass index; WC, waist circumference; SBP, systolic blood pressure; DBP, diastolic blood pressure; MAP, mean arterial pressure; BDNF, brain-derived neurotrophic factor.

**Table S4. Adverse events during the eBACH trial.**

| Adverse Event | Treatment Group | Days on Intervention | Reason | Relationship to Intervention | Outcome |
|---|---|---|---|---|---|
| 1 | Aerobic Exercise | 28 | Ankle sprain | No | Recovered, with treatment |
| 2 | Aerobic Exercise | 55 | Back Pain | No | Recovered, with treatment |
| 3 | Aerobic Exercise | 221 | Back Pain | No | Recovered, with treatment |
| 4 | Aerobic Exercise | 298 | Bone fracture | No | Recovered, with treatment |
| 5 | Aerobic Exercise | 41 | Cardiovascular | No | Still present, being treated |
| 6 | Aerobic Exercise | 11 | Foot fracture | No | Recovered, with treatment |
| 7 | Aerobic Exercise | 66 | Removal of tumor on parotid gland | No | Recovered, with treatment |
